# Digitizing Non-Invasive Neuromodulation Trials: Scoping Review, Process Mapping, and Recommendations from a Delphi Panel

**DOI:** 10.1101/2022.03.03.22271837

**Authors:** Andre R Brunoni, Hamed Ekhtiari, Andrea Antal, Paradee Auvichayapat, Chris Baeken, Isabela M. Benseñor, Marom Bikson, Paulo Boggio, Barbara Borroni, Filippo Brighina, Jerome Brunelin, Sandra Carvalho, Wolnei Caumo, Patrick Ciechanski, Leigh Charvet, Vincent P. Clark, Roi Cohen Kadosh, Maria Cotelli, Abhishek Datta, Zhi-De Deng, Rudi De Raedt, Dirk De Ridder, Paul B. Fitzgerald, Agnes Floel, Flavio Frohlich, Mark S. George, Peyman Ghobadi-Azbari, Stephan Goerigk, Roy H. Hamilton, Shapour J. Jaberzadeh, Kate Hoy, Dawson J. Kidgell, Arash Khojasteh Zonoozi, Adam Kirton, Steven Laureys, Michal Lavidor, Kiwon Lee, Jorge Leite, Sarah H. Lisanby, Colleen Loo, Donel M. Martin, Carlo Miniussi, Marine Mondino, Katia Monte-Silva, Leon Morales-Quezada, Michael A. Nitsche, Alexandre H. Okano, Claudia S. Oliveira, Balder Onarheim, Kevin Pacheco-Barrios, Frank Padberg, Ester M Nakamura-Palacios, Ulrich Palm, Walter Paulus, Christian Plewnia, Alberto Priori, Tarek K. Rajji, Lais B. Razza, Erik M. Rehn, Giulio Ruffini, Klaus Schellhorn, Mehran Zare-Bidoky, Marcel Simis, Pawel Skorupinski, Paulo Suen, Aurore Thibaut, Leandro C. L. Valiengo, Marie-Anne Vanderhasselt, Sven Vanneste, Ganesan Venkatasubramanian, Ines R. Violante, Anna Wexler, Adam J. Woods, Felipe Fregni

## Abstract

Although relatively costly and non-scalable, non-invasive neuromodulation interventions are treatment alternatives for neuropsychiatric disorders. The recent developments of highly-deployable transcranial electric stimulation (tES) systems, combined with mobile-Health technologies, could be incorporated in digital trials to overcome methodological barriers and increase equity of access. We convened 61 highly-productive specialists and contacted 8 tES companies to assess 71 issues related to tES digitalization readiness, and processes, barriers, advantages, and opportunities for implementing tES digital trials. Delphi-based recommendations (>60% agreement) were provided. Device appraisal showed moderate digitalization readiness, with high safety and the possibility of trial implementation, but low connectivity. Panelists recognized the potential of tES for scalability, generalizability, and leverage of digital trials processes; although they reached no consensus about aspects regarding methodological biases. We further propose and discuss a conceptual framework for exploiting shared aspects between mobile-Health tES technologies with digital trials methodology to drive future efforts for digitizing tES trials.

**Graphical Abstract. Consensus Roadmap:** 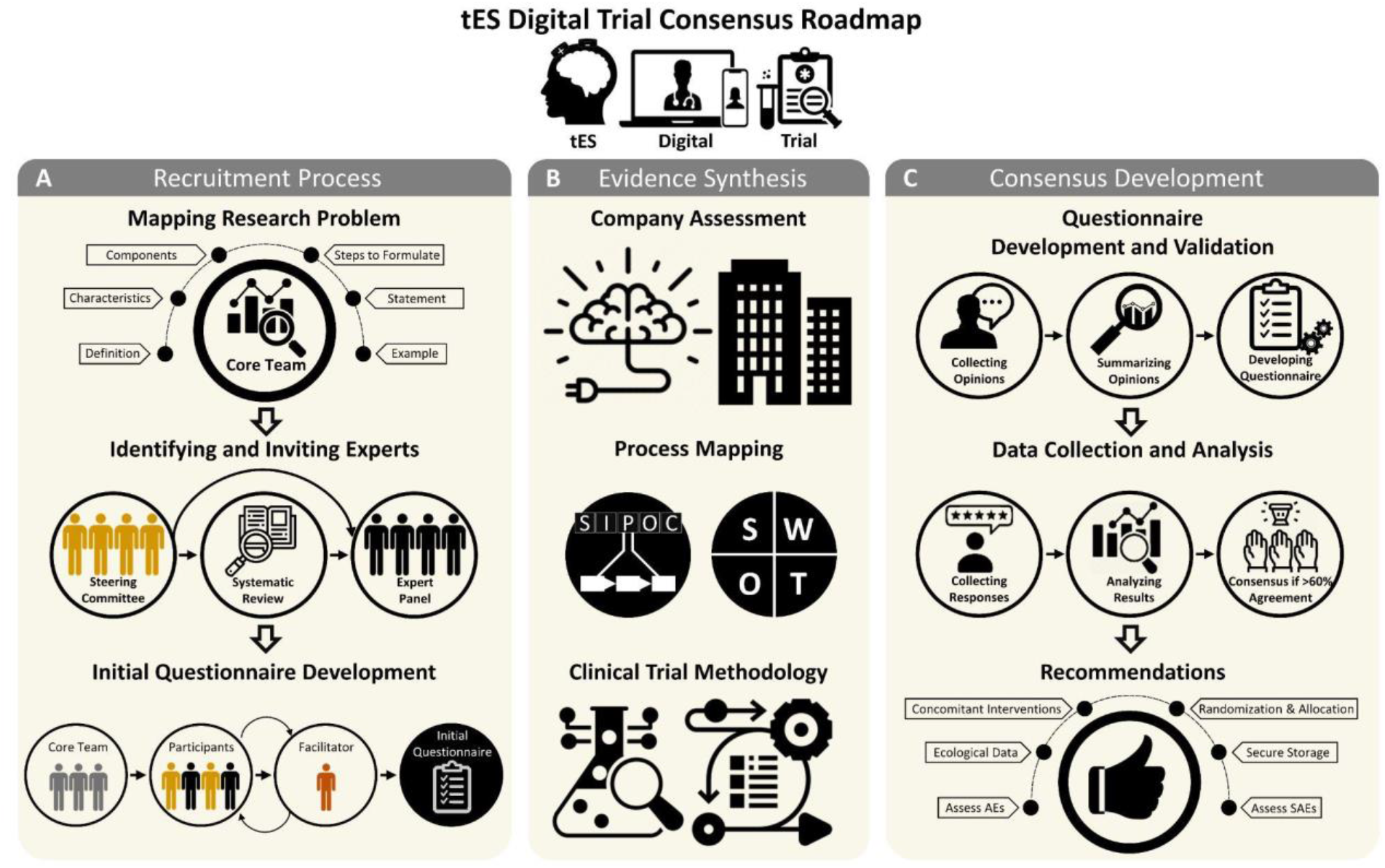

*(A) Recruitment process*. The study procedure started with defining the components of the research problem by the core research team. After defining the problems, two different sets of participants (the steering committee (SC) including key leaders of the field identified by the core team and the expert panel (EP) as a more diverse group of experts identified based on the number of publications based on a systematic review) were identified and were invited to participate in a Delphi study. The study facilitators (first and last authors) led the communications with the SC to design the initial questionnaire through an iterative approach. *(B) Evidence synthesis:* To collect the available evidence, companies producing portable tES (ptES) devices were contacted, based on the companies suggested by the SC and EP to provide details about the available devices. For mapping methodological processes of digitizing tES trials, two distinct strategies were performed and embedded into the questionnaire, namely SIPOC (Suppliers, Inputs, Process, Outputs, and Customer) and SWOT (Strengths, Weaknesses, Opportunities, and Threats) assessment were performed and embedded into the questionnaire. *(C) Consensus development:* In the next phase, the questionnaire was validated and finalized via collecting and summarizing opinions. Afterward, the SC and EP responded to the final questionnaire, and results were analyzed providing a list of recommendations for running tES digital trials based on a pre-registered consensus threshold.

## 1. Introduction

Transcranial electric stimulation (tES) is a non-invasive neuromodulation intervention that uses electric currents applied over the scalp to modify cortical activity and treat neuropsychiatric disorders and has high safety and tolerability (Fregni et al., 2021). Notwithstanding, due to its low-to-moderate efficacy for several conditions, the consensus of its readiness for clinical use across indications varies (Ekhtiari et al., 2019; Fregni et al., 2021), and regulatory approvals across regions are mixed (Bikson et al., 2018b; Fregni et al., 2015), warranting larger-scale clinical trials (Brunoni et al., 2012). However, these trials are hampered by the need for daily visits to the research center to deliver the necessary number of tES sessions, limiting recruitment, harming adherence, increasing costs (5), and restricting diversity (Bikson et al., 2018a; Charvet et al., 2020).

Unlike other non-invasive neuromodulation modalities, tES devices, by virtue of being affordable and battery-powered (Woods et al., 2016), are portable, intervening an appealing brain stimulation modality for patients who do not tolerate pharmacotherapy (Brunoni et al., 2019) or have difficulty attending treatment at a clinical center several companies have been designing highly-deployable tES devices that could be used to address issues of scale, access, and patients’ burden in the context of digital trials - i.e., trials that leverage aspects such as recruitment, assessment, and data analyses through the implementation of digital technologies (Inan et al., 2020). These approaches could be further integrated with mobile Health devices, apps, and wearables, allowing for several new implementations, such as simultaneous combination with cognitive and psychological interventions, ecological momentary assessment of behaviors, passive data collection, and digital phenotyping (Insel, 2018; Torous et al., 2017). However, since protocols and standards for digital trials using mobile tES are still emerging, the challenges and opportunities of their implementation processes have not yet been systematically examined. Moreover, issues on rigor and reproducibility - for instance, best practices to perform randomization, allocation concealment, and sham stimulation - and generalizability - how to fully explore their potential for scalability while ensuring adherence and representativeness - have only been discussed in non-digital contexts (Bikson et al., 2018a; Charvet et al., 2020, 2015). Finally, implementation challenges are different in low-/middle-income countries, which present lower digital literacy, fluency in non-native languages, and wireless connectivity (Silva-Filho et al., 2022); conversely, scalable mobile Health interventions have even higher impact potential in such countries (Torous et al., 2021).

Considering these challenges, we systematically identified non-invasive neuromodulation specialists to elaborate and discuss issues related to the extent, processes, and methodological characteristics of digitizing non-invasive neuromodulation trials. These findings, supplemented by a systematic scoping review of tES clinical research articles and an assessment of the digitalization degree of commercially available tES devices, provided a key summary of Delphi-based recommendations for enhancing the implementation of digital tES trials.

## 2. Methods

Our protocol was pre-registered at the Open Science Framework (OSF) (Razza et al., 2021) and is depicted in Fig. 1. Minor deviations occurred (Sup. Material - Appendix 1).

**Fig. 1.**
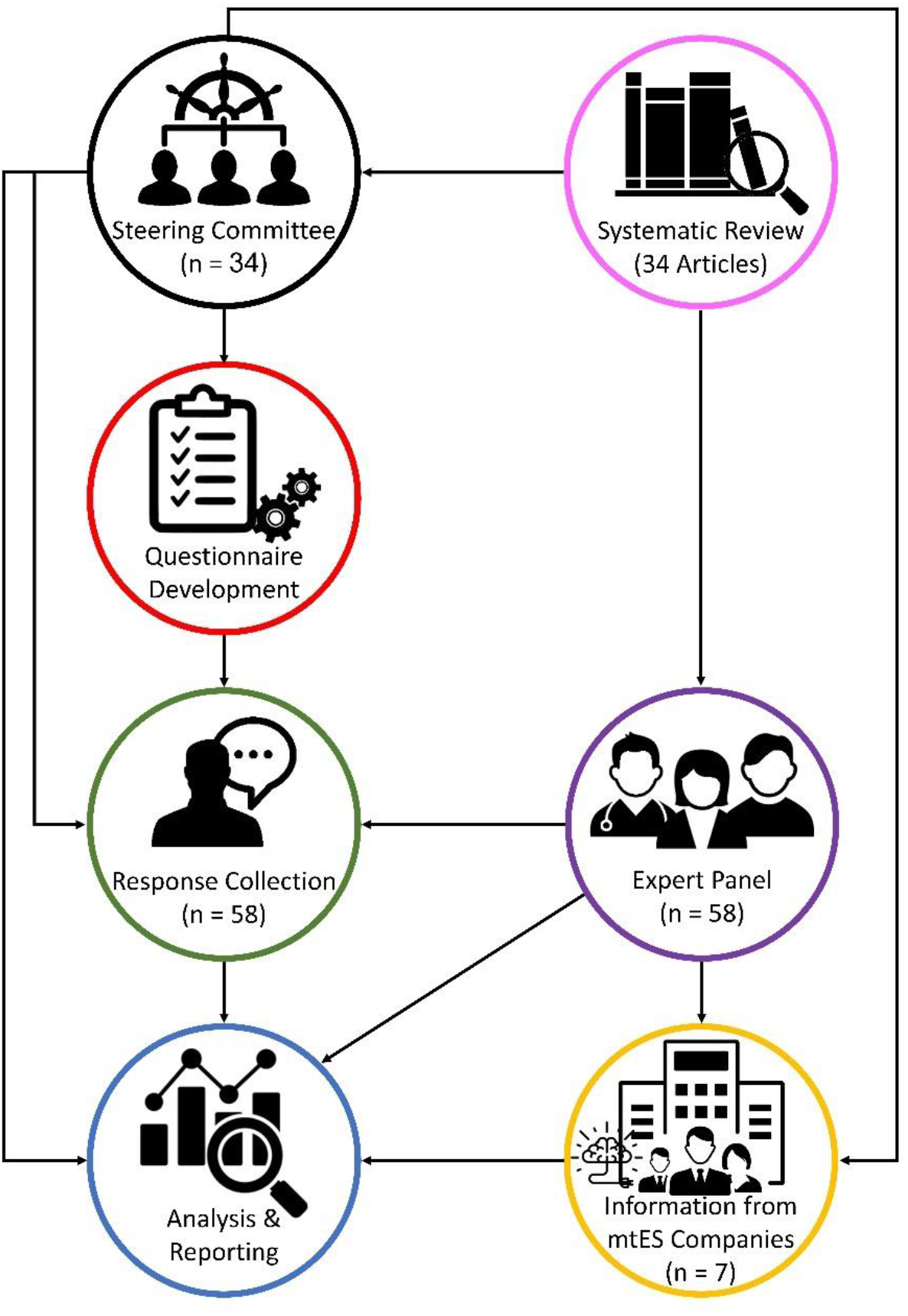
Study workflow. First, the steering committee (SC) was formed, including prominent researchers in the field. Then, supplemented by the results of a systematic review conducted on tES trials, the SC developed the questionnaire, which was sent to all participants of the study (SC and expert panel (EP)) to answer it. Simultaneously, companies producing tES devices were contacted, based on the companies known by the SC and EP, to provide details about the available devices. Finally, the SC analyzed the data received from the questionnaire and all participants took part in reporting the results.

### 2.1. Specialist panel

We used a Delphi technique, in which comments and feedback are iteratively discussed (Hsu and Sandford, 2019), for addressing challenges and proposing recommendations for digitizing tES trials. Following recent papers (Ekhtiari et al., 2022a, 2022b), we initially convened a steering committee (SC) group, formed based on the collaborative network of the leading authors, to develop structured questionnaires with items using Five-point Likert scales or open questions (Supplementary Material - Appendix 2). The SC also provided qualitative feedback on several topics that were analyzed by the leading authors and qualitatively described here. Afterward, the SC interacted with a larger expert panel (EP) to rate each item. The EP members were selected among the most productive authors in the field, identified through a systematic review of recent tES clinical trials in 10 years (Sup. Material - Appendix 3). Several interactions were performed between the EP and SC until a final manuscript version was assembled. The consensus was achieved by a >60% agreement of all panelists. Electronic questionnaires were used in all steps of this process. All members of the SC and the EP consented to have their names listed and identified in the manuscript.

### 2.2. Systematic scoping review

A systematic scoping review (Levac et al., 2010) was performed to identify tES reviews, consensus papers, and guidelines to select characteristics for composing the questionnaires used in the rating phase (Sup. Material - Appendix 4).

### 2.3. tES digitalization readiness

Companies producing tES devices were identified through specialist referrals and web search and surveyed using structured questionnaires to assess their digitalization readiness, according to connectivity, readiness for digital trials, parameter space flexibility, ecological footprint, front-end interface, and data security (Sup. Material - Appendix 5).

### 2.4. Process mapping and methodological assessment

We used SWOT (Strengths, Weaknesses, Opportunities, and Threats) and SIPOC (Suppliers, Inputs, Process, Outputs, Customers) approaches to respectively identify external and internal negative and positive aspects for digitizing tES trials and map and compare processes related to standard and digital tES trials. The methodological assessment was based on perceived challenges and advantages, identified through questionnaires, of conducting such trials (Sup. Material - Appendix 2).

## 3. Results

### 3.1. Specialist panel

For the SC, 34 authors were invited and all agreed to participate. For the EP, out of 43 authors who were identified, 14 did not reply to our contacts, and 2 declined to participate. Finally, 27 participants constituted the EP (Sup. Material - Appendix 6). Most panelists were men (70%), experienced (78% with > 10 years of experience in the field), and between 40 to 49 years old (44% and 33% of the SC and the EP). They resided in the US (SC n=11, EP n=3), Brazil (SC n=6, EP n=5), Germany (SC n=5, EP n=4), and 13 other countries (Sup. Material - Appendix 7). Only 15% and 18% of the SC and EP members, respectively, were not conducting at least one tES trial with digital features; most were principal investigators (83%) of such trials.

### 3.2. Systematic scoping review

Our initial search yielded 443 references, and 34 articles met the inclusion criteria of our scoping review, including 9 recommendation articles (Bikson et al., 2018a; Deer et al., 2014; Fregni et al., 2015; Fried et al., 2021; Kim et al., 2020; Sandars et al., 2016; Sierawska et al., 2019; Thibaut et al., 2017; Zhang et al., 2019), 10 guidelines (Antal et al., 2017; Bikson et al., 2020; Charvet et al., 2020, 2015; Cruccu et al., 2016; Fregni et al., 2021; Gillick et al., 2018; Lefaucheur et al., 2017; Legatt et al., 2016; Parikh et al., 2016), 10 critical reviews (ALHarbi et al., 2017; Cappon et al., 2016; Godeiro et al., 2021; Lucchiari et al., 2018; Maatoug et al., 2021; McClintock et al., 2019; Sanches et al., 2021; Santos et al., 2021; Shiozawa et al., 2017; Workman et al., 2020), and 5 expert consensus articles (Baptista et al., 2019; Buch et al., 2017; Ekhtiari et al., 2019; Grimaldi et al., 2014; Martelletti et al., 2013), which were used for elaborating the study questionnaires (Sup. Material - Appendix 8).

### 3.3. tES digitalization readiness

Eight of 13 companies contacted provided feedback. Digitization readiness varied according to their wireless connectivity, readiness for digital trials, the flexibility of parameter space, ecological footprint, front-end interface, and data security. Markedly, current systems have limited wireless connectivity, which is a barrier for device-to-device communication with wearables and third-party apps that could enhance portability potential (e.g., apps collecting biological data, and mobile mental health apps). Conversely, most systems currently present good data security protocols (reported HIPAA or GDPR compliance), the flexibility of tES parameter space, and readiness for digital trials (Fig. 2).

**Fig. 2.**
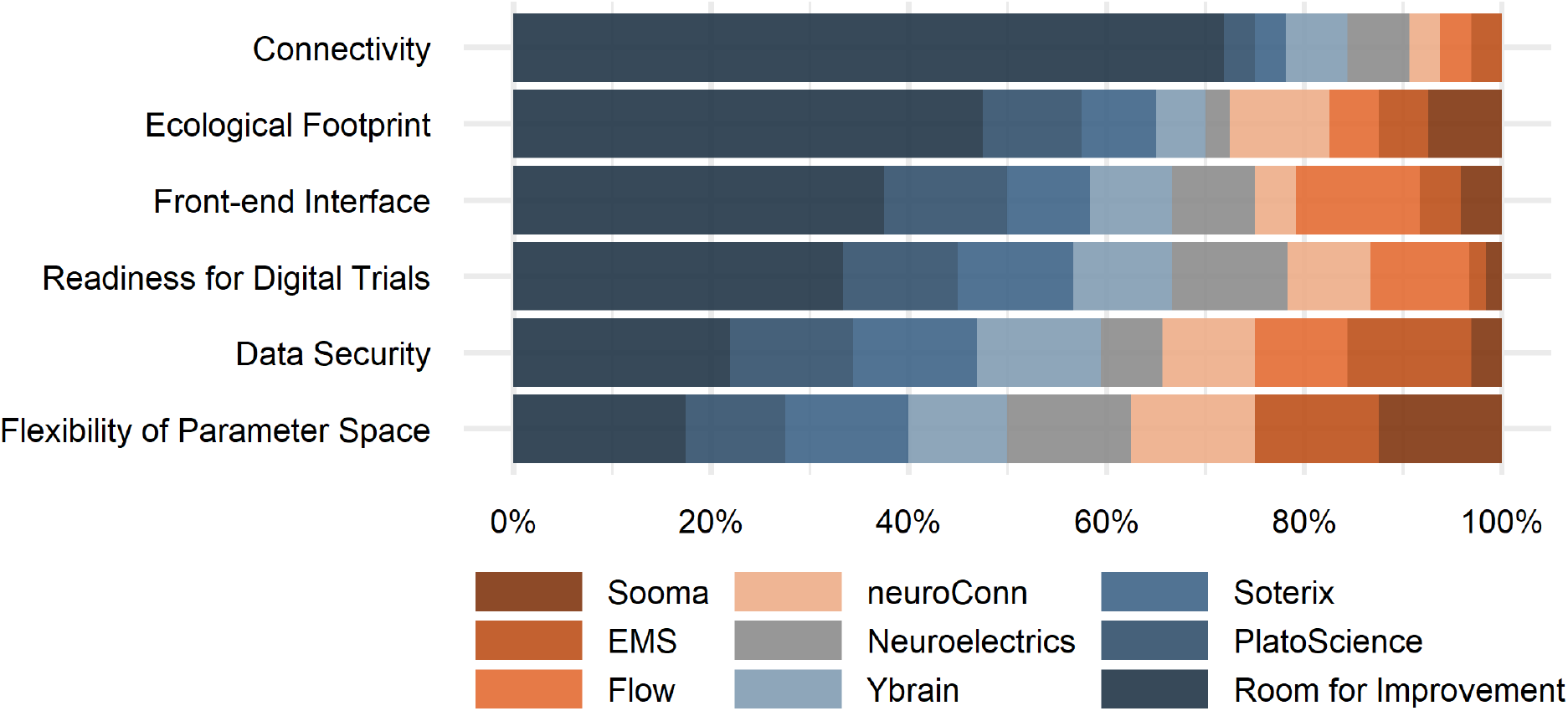
Digitalization readiness of tES devices. Based on the feedback of 8 out of 13 major transcranial electric current stimulation (tES) companies, we evaluated the readiness of these systems for digital trials, considering their connectivity (capability of communicating with other devices and the Web, due to the presence of Bluetooth, Wi-Fi, 3G/4G/5G, and communication with third-party apps), methodological aspects (randomization, sham, blinding, built-in data collection, optional data collection, and optional research dashboard), parameter space flexibility (current intensity, session duration, number of sessions, electrode positioning), ecological footprint (rechargeability and replaceability of batteries, recyclability and reusability of sponges, and recyclability of devices), front-end interface (smartphone app, touch screen, device itself, no interaction), and data security (compliance to laws such as GDPR, mention of encryption and anonymization procedures, and option for not collecting sensitive data). The questionnaires and rating systems are described in the Sup. Material-Appendix 5.

### 3.4. SWOT assessment

The identified characteristics and quantitative agreement rating composing the SWOT assessment are displayed in Fig. 3. Qualitative aspects are briefly discussed here and detailed in the Sup. Material - Appendix 9.

**Fig. 3.**
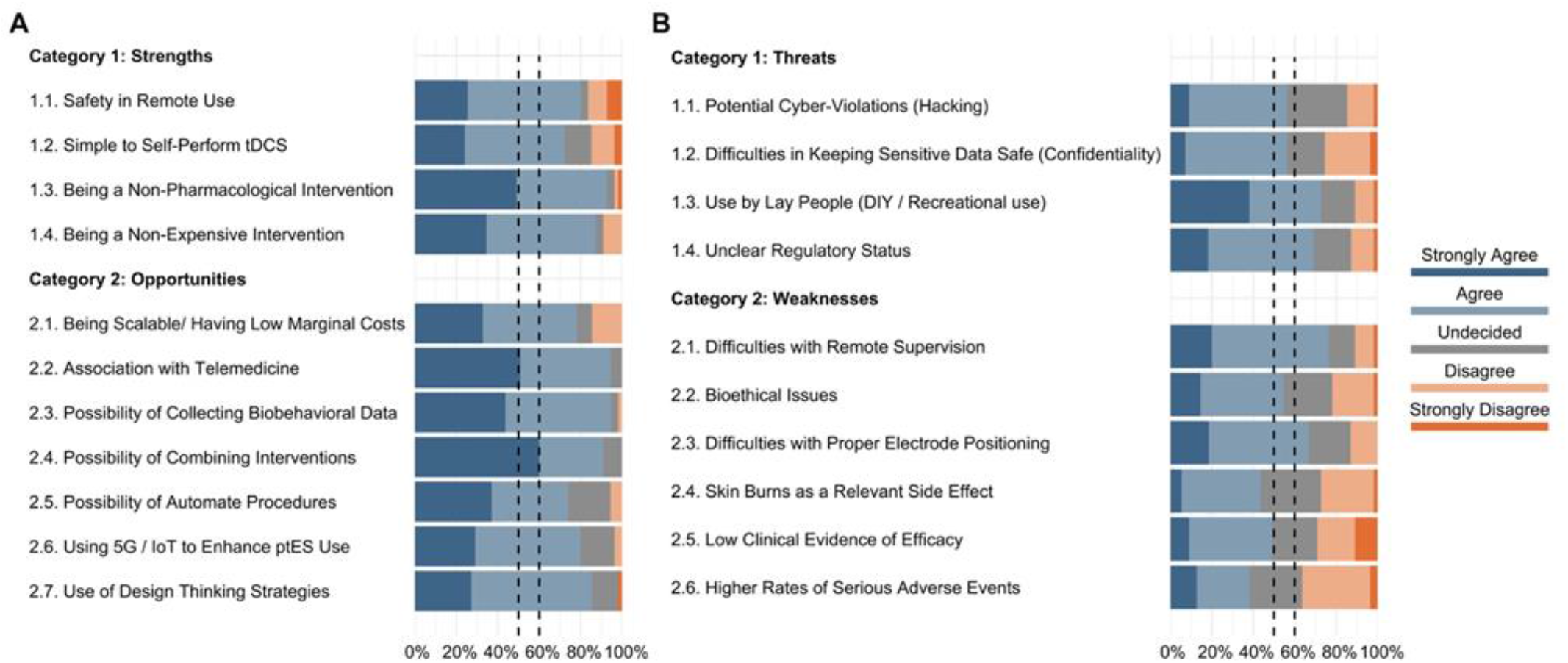
SWOT (Strengths, Weaknesses, Opportunities, and Threats) assessment for digital tES trials. This figure depicts the ratings of 55 participants (24 from the steering committee and 31 from the expert panel) for the ptES clinical trials strengths and opportunities (A) and threats and weaknesses (B). Each item was rated from strongly disagree to strongly agree. In ratings for the potential strengths and opportunities of tES clinical trials (A), all of the items have reached the 50% threshold of agreement (rated as either agree or strongly agree by more than 50% of the respondents). These items have also reached a more stringent threshold of 60%. However, in ratings for the potential threats and weaknesses of ptES clinical trials (B), two of the items (2.4. Skin burns as a relevant side effect, and 2.6. Higher rates of serious adverse events) did not reach the 50% threshold. Items are represented by their summary in the figure. Full text of the items is provided in Supplementary Tables 1 and 2. tDCS=transcranial direct current stimulation. ptES=portable transcranial electrical stimulation. DIY=do-it-yourself.

Regarding strengths, the panelists agreed on four features: (a) high safety, considering previous evidence from non-digital trials and studies in humans (Antal et al., 2017; Aparício et al., 2016; Bikson et al., 2016; Moffa et al., 2017); (b) feasibility of self-application, owing to recent developments of devices in which electrode placement is fixed, methods for easy strap positioning, and friendly end-user interface of mobile tES device (Charvet et al., 2020); (c) being a non-pharmacological intervention; and (d) affordability, as tES devices are simple to be built in terms of electric engineering (Woods et al., 2016), costs of high-end features (e.g., microprocessors, Bluetooth and wireless connectivity, miniaturization) are decreasing over time, and self-application saves human resources.

**Table 1.** Comparison of SIPOC processes of trials in which digital features are present and absent.

| Digital features | Suppliers | Inputs | Outputs | Clients |
| --- | --- | --- | --- | --- |
| **Process: Recruitment** |  |  |  |  |
| *Absent* | Traditional Media | Telephone call or email | Pool of volunteers constrained due to geographical barriers | Pre-screening / screening onsite teams |
| *Present* | Targeted Social Media and Google Ads, public segmentation | Electronic forms, ChatBots | Larger pool of volunteers | Online pre-screening / screening teams |
| **Process: Pre-screening** |  |  |  |  |
| *Absent* | Volunteers | Onsite checklist of eligibility criteria | Potential Participants | Onsite screening team |
| *Present* | Volunteers | Online assessments, AI techniques can increase the likelihood of inclusion | Potential Participants | Online screening team |
| **Process: Screening** |  |  |  |  |
| *Absent* | Potential participants | Onsite clinical interview; written consent | Participants | Onsite clinical team |
| *Present* | Potential participants | Clinical interviews aided by digital assessments; digital consent | Participants | Online clinical team |
| **Process: Participation in the trial** |  |  |  |  |
| *Absent* | Participants | Staff delivers tES sessions and assessments | Completers and patients who dropped out | Trial complete; possibly follow-up studies |
| *Present* | Participants | tES devices shipped to participants; online videos and telesupport to guide self-applied tES; digital assessments via online interviews and mHealth technologies | Completers and patients who dropped out | Trial complete; possibly follow-up studies |

The table illustrates how processes of clinical trials can be leveraged using digital approaches. In (a) recruitment, volunteers, unconstrained by geographical barriers, can fill out electronic forms and be automatically excluded according to eligibility criteria; (b) pre-screening, potential participants can be scheduled and contacted automatically for online screening (a step that can be enriched by using machine learning algorithms); (c) screening, enrolled participants can provide digital consent, have transcranial electric stimulation (tES) devices shipped to their homes, and being instructed how to use them via videos and/or augmented reality techniques; and (d) participation in the trial, interaction with the staff can be mediated by psychiatric chatbots for filtering between simple questions, reporting of adverse events and need of medical care.

Regarding weaknesses, panelists agreed on two aspects: (a) difficulties in remote supervision, raising concerns regarding patients themselves manipulating tES devices, which could lead to misuse, diversion of the device, or its use outside of medical contexts, further impacting on the reproducibility of findings; (b) and difficulties in obtaining accurate placement of electrodes, as deviations in electrode positioning and orientation might affect outcomes (Woods et al., 2016). Therefore, companies should develop and test new methods for assuring the correct placement of electrodes (Fried et al., 2021). Other potential weaknesses did not reach consensus, such as concerns related to bioethics, particularly regarding equity to access; increased (compared to on-site tES trials) risks of common and serious adverse events (Lefaucheur et al., 2017); and relatively low evidence of clinical efficacy for most conditions (Fregni et al., 2021).

Regarding opportunities, six aspects reached agreement: (a) scalability, as, compared to on-site tES trials that need physical space, staff to apply sessions and commute of participants, digital trials using mobile tES devices do not have such constraints, allowing research assistants to monitor several participants at once, at any distance from the study centers; (b) telemedicine, which has been widely adopted during the COVID-19 pandemic, facilitating its adoption in digital trials; (c) employment of combined mobile Health technologies, permitting digital phenotyping (Torous et al., 2017) and combination with digital interventions when using paired wearables and smartphone applications; (d) automatization of procedures (see SIPOC below); (e) 5G / Internet of Things, which can boost connectivity and data processing, leveraging data collection (Torous et al., 2017) and eventually allowing the development of mobile closed-loop tES systems (Sanches et al., 2021); and (f) use of design thinking approaches, i.e., customizing mobile tES devices around the patients’ perspectives (Polhemus et al., 2020), for instance, to accommodate those with physical or cognitive impairments.

Finally, two threats reached a consensus: (a) recreation and do-it-yourself misuse, which could lead to unexpected adverse events and safety issues (Sierawska et al., 2019); and (b) regulatory status, as medical devices require formal regulatory approval in the US (Darrow et al., 2021) and Europe (Antich-Isern et al., 2021), although some tES devices are marketed as wellness devices, have regulatory device exemptions (Bikson et al., 2018b), or can be approved by similarity (Bikson et al., 2018b). Further, mobile tES devices could have additional regulations, if framed as mobile Health systems (Onodera and Sengoku, 2018). Additionally, two potential threats were identified by most of the panel, but did not reach the 60% consensus threshold: (a) risks related to hacking and cyber-security, as observed in mobile Health devices (Aljedaani and Babar, 2021), and (b) risks related to confidentiality and anonymity.

### 3.5. SIPOC

We identified 4 main processes (recruitment, pre-screening, screening, and participation) in which digitization and automatization procedures can leverage mobile tES trials (Table 1). The panelists noted that trials might not be purely digital or analog, and different degrees of digital features can occur at each process. For instance, participants can be recruited through social media but screened onsite. Moreover, digital processes can provide enhanced metrics to adjust processes, recruit faster, and follow participants for longer periods. Finally, digitization processes provide scalability due to the use of digital assessments and telemedicine. (Table 1).

### 3.6. Methodological aspects

The panelists examined 24 methodological aspects of digital trials using mobile tES devices, reaching consensus in 12 of them (Fig. 4). They are briefly discussed here and detailed in the Sup. Material - Appendix 10.

**Fig. 4.**
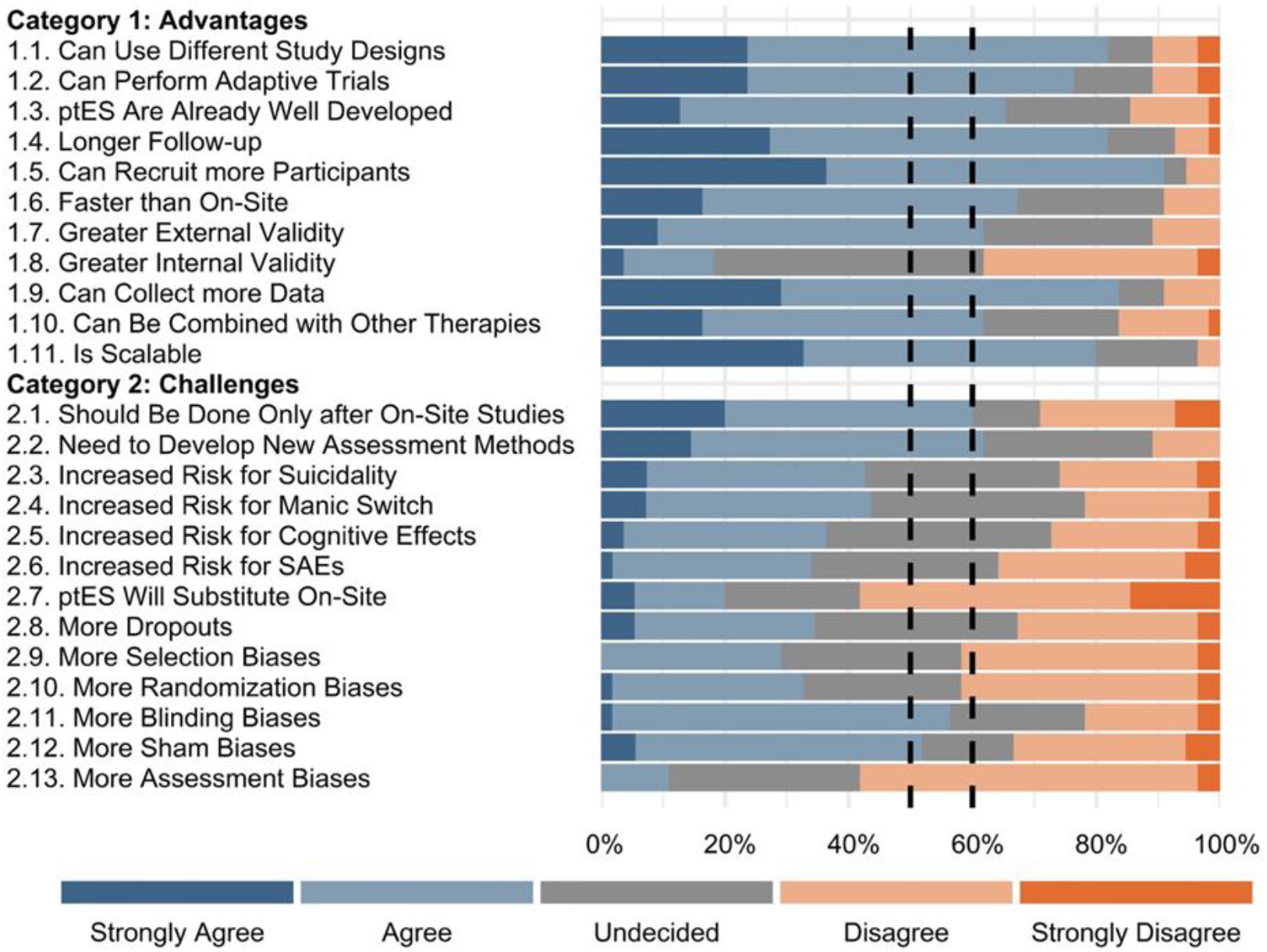
Ratings for advantages and challenges of conducting tES digital trials. This figure depicts the ratings of 55 raters (24 from the steering committee and 31 from the expert panel) for methodological aspects of tES. Each item was rated from strongly disagree to strongly agree. In ratings for the advantages of transcranial electric stimulation (tES) digital trials from a methodological perspective, only one item (1.8. Greater internal validity) did not reach the agreement threshold of 50% (rated as either agree or strongly agree by more than 50% of the respondents). All of the other items in this category also reached a more stringent threshold of 60%. However, in ratings for challenges of tES digital trials from a methodological perspective, 9 items (2.3. Increased risk for suicidality, 2.4. Increased risk for the manic switch, 2.5. Increased risk for cognitive effects, 2.6. Increased risk for SAEs, 2.7. ptES will substitute on-site, 2.8. More dropouts, 2.9. More selection biases, 2.10. More randomization biases, and 2.13. More assessment biases) did not reach the 50% threshold of agreement. With a more stringent threshold of 60%, two additional items (2.11. More blinding biases and 2.12. More sham biases) dropped out from the agreement. Items are represented by their summary in the figure. Full texts of the items are provided in Supplementary Table 3. ptES=portable transcranial electrical stimulation. SAE=serious adverse event.

Of the 12 aspects that reached consensus, 10 were perceived as advantages of digital trials, which included (a) the adoption of different study designs, including (b) adaptive designs, as adaptation rules can be performed automatically and remotely whether data are collected and analyzed by mobile tES systems. Panelists also considered that (c) tES devices are already sufficiently developed to be used remotely, which allows for (d) longer follow-up periods and (e) higher recruitment rates, being (f) faster and more efficient than on-site trials. Also, (g) greater external validity compared to on-site trials were perceived. Finally, other advantages were the potential for (h) collecting more data than on-site trials, (i) combination with other therapies and (j) scalability. The 2 disadvantages/challenges that reached consensus were: (a) the necessity of validating new tES parameters, methodologies, and indications first in on-site studies, and (b) the need of developing better remote assessment methods, such as behavioral clinical scales properly designed and validated to be employed remotely.

The items that did not reach consensus mostly pertain to internal validity issues. Interestingly, a significant portion of panelists was undecided on these issues (Fig. 5). Of note, interesting remarks (detailed in Sup. Material - Appendix 10) were made for: (a) randomization-allocation procedures that are done using either specific devices or apps/software, but can be vulnerable to contamination biases due to hacking and exposure of the randomization list; (b) study blinding, as blinding breaking can occur if devices are manipulated; (c) sham stimulation, which can also be revealed due to device manipulation.

**Fig. 5.**
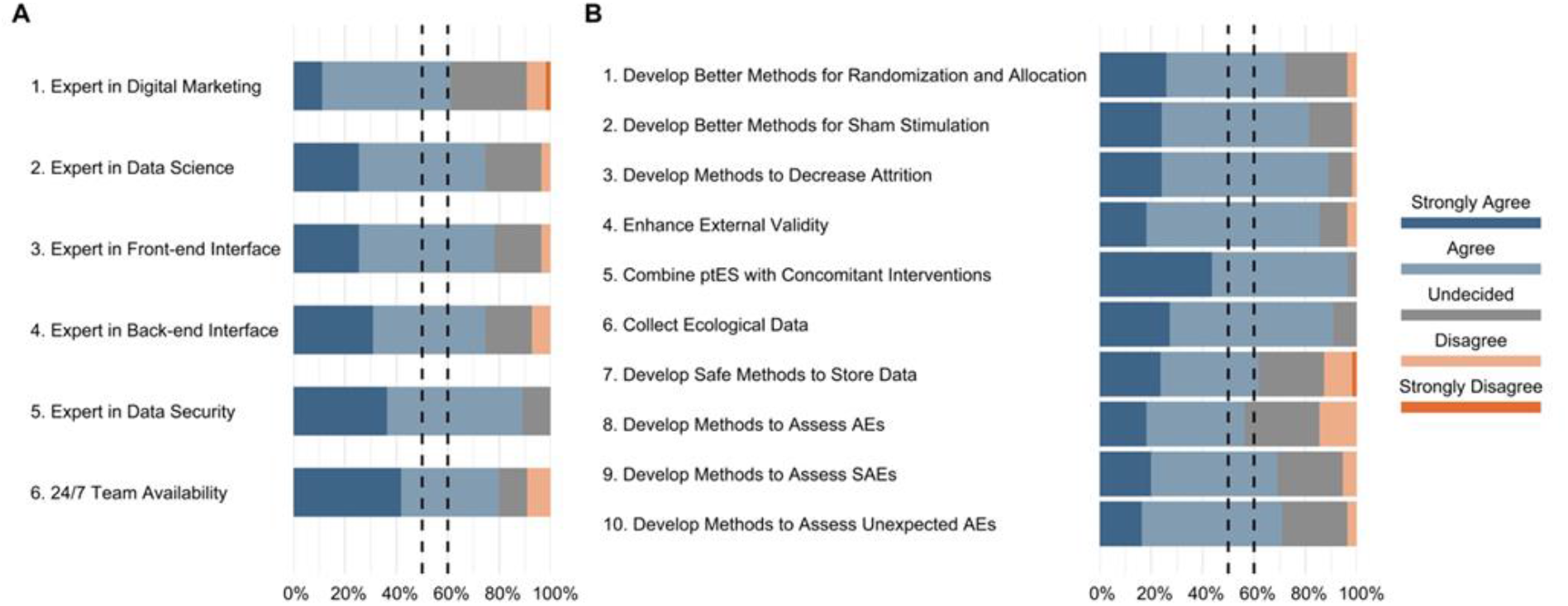
Ratings of the tES clinical trials team features and general recommendations on digitizing tES trials. This figure depicts the ratings of 55 participants (24 from the steering committee and 31 from the expert panel) for the ptES clinical trials team features (A) and general recommendations for digitizing ptES clinical trials (B). Each item was rated from strongly disagree to strongly agree. In ratings for the ptES clinical trials team features (A), all of the items have reached the 50% threshold of agreement (rated as either agree or strongly agree by more than 50% of the respondents). These items have also reached a more stringent threshold of 60%. Similarly, in ratings for the general recommendations for digitizing ptES clinical trials (B), all of the items have reached the 50% threshold of agreement. However, with a more stringent threshold of 60%, one item (8. Develop methods to assess AEs) dropped out from the agreement. Items are represented by their summary in the figure. Full text of the items is provided in Supplementary Tables 4 and 5. ptES=portable transcranial electrical stimulation. AE=adverse event. SAE=serious adverse event.

### 3.7. Recommendations

The panelists recommended that teams performing tES digital trials should have members specialized in (a) digital marketing strategies, to enhance online recruitment; (b) data science and visualization, for data collection and analysis; (c) front-end interfaces, to enhance user experience; (d) back-end programming; (e) issues related to data security, integrity, anonymity, and replicability. Also, they suggested that (f) a team member should be always (“24/7”) available (Fig. 5).

Regarding further research, most panelists recommended that better methods for (a) randomization/allocation, and (b) sham should be explored. Also, further research was recommended to develop or refine methods to enhance (c) adherence and (d) external validity of the trials. Also, more research should be devoted to aspects such as (e) combination of interventions, (f) biobehavioral data collection, (g) enhanced data security, and methods to assess (h) serious and (i) unexpected adverse events (details in Sup. Material - Appendix 11).

## 4. Discussion

By convening a diverse group of 61 worldwide specialists in the field of non-invasive neuromodulation, we performed the first systematic assessment and Delphi-based validation of perceived challenges, opportunities, methodological issues, and recommendations on digitizing non-invasive neuromodulation trials. We used several strategies to organize and describe these assessments, such as processing mapping strategies, a systematic scoping review of the literature, iterative rating and validation of structured questionnaires by specialists, and assessment of the digital readiness of commercial tES devices. Taken together, our findings show that performing digital trials using mobile tES devices has complementary advantages and can overcome major on-site tES trial challenges, namely the intensive treatment schedules (Thibaut et al., 2017), transportation costs, accessibility, and scalability (Charvet et al., 2020). By performing trials remotely, dislocation burdens are decreased, as well as the need for space at the research center, and of trained staff for delivering tES sessions, aspects that increase the trial duration (Brunoni et al., 2012; Parikh et al., 2016, p. 4). Additionally, tES devices are highly scalable, as a single team member can monitor several people at once, provided that tES devices are easy to manipulate, handle, and can be self-delivered. Such scalability gains could be leveraged in faster trials with larger sample sizes, longer follow-up periods, and employing digital recruitment strategies. Considering our results, we propose and further discuss a conceptual framework for digitizing neuromodulation, combining concepts of digital clinical trials with mobile tES (Fig. 6).

**Fig. 6.**
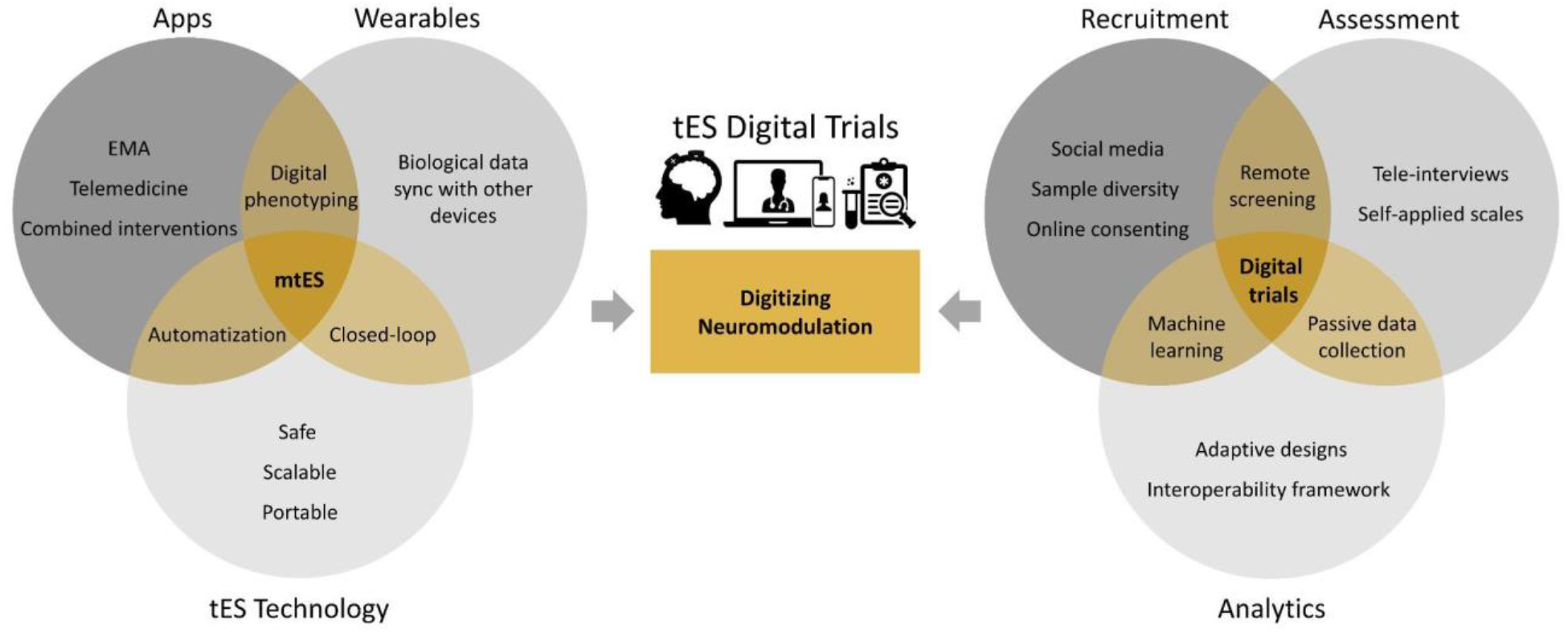
A conceptual framework for digitizing neuromodulation. As depicted on the left side of the figure, transcranial electric stimulation devices had been relatively simple, essentially using batteries connected to electrodes to deliver constant currents, and contain few (micro) electronic components. Although portable and safe, they had not been specifically designed for use outside hospital or academic center settings. New and future generations of tES will be mobile Health tES systems using digital technologies for improving health outcomes. They are and will be smaller and lighter than previous generations, possessing wireless connectivity. Such devices are already used at home and are self-delivered, usually with some degree of remote supervision. Their use will be supported by proprietary or third-party apps and wearables. Resulting together with the aforementioned concept as digitizing neuromodulation, the right side of the figure shows digital trials as clinical trials that use digital features to enhance recruitment, assessment, and data analysis and could unleash the full potential of tES regarding scalability and equity of access. There are many similarities between the assumptions of digital trials and the capabilities of mtES, which are discussed in this work. EMA=Ecological momentary assessment. mtES=mobile transcranial electrical stimulation.

### 4.1. Mobile tES devices

To the best of our knowledge, the term “mobile tES” has not been used yet to describe the combination of highly-deployable neuromodulation devices paired with other wearables or apps. This terminology frames tES in the context of mobile Health devices (Onodera and Sengoku, 2018), encouraging the exploitation of contact points between these two growing fields. However, even though several issues for deployable and remote use have already been addressed (e.g., decreasing prices, rechargeable batteries, tailored sponges, sham stimulation, easiness of electrode positioning, and programming session stimulation parameters), our findings showed that no commercial tES devices have been fully digitized yet, presenting different degrees of online, wired, or wireless connectivity. Also, especially for offline devices, methods for restricting the number of sessions allowed per day were not identified. Additionally, most systems neither collect active or passive data, nor present friendly-user interfaces.

The panelists agreed on several opportunities; however, most are distant from immediate implementation. For instance, device-to-device communication (“Internet of Things”) would need pairing with third-party apps or wearables, which is not yet available. This limits other perceived opportunities such as digital phenotyping, combination with psychological or cognitive app interventions, and seamless automatization with other platforms and digital processes.

### 4.2. Methodological challenges, advantages, and processes of digital trials

The impact of digitizing neuromodulation trials on external validity seems mostly positive, considering that subjects who would not be enrolled in on-site trials are reached. On-site trial samples are likely to be composed of those with free time and/or living near the clinical center to receive the sessions. Notwithstanding, it is still possible that those younger, richer, more educated, with higher digital literacy, and living in urban areas are preferentially enrolled in digital trials. Also, if recruitment strategies are performed solely using social media, the trial results would have restricted generalizability for people that do not use such media. This could be overcome by using segmented digital marketing campaigns. Likewise, attrition rates might not necessarily be lower in digital trials - although not needing to return daily to the clinical center could decrease the burden and minimize dropouts, samples from digital trials might face less engagement and more difficulties in self-delivering the sessions. The lack of daily contact with the clinical staff could also decrease motivation and increase dropouts. In addition, direct social contact, social support, and social connectedness outside the digital environment can influence attrition rates of clinical trials. Therefore, telemonitoring and proper interaction with participants, managing their expectations and credibility of the team, and reinforcing the need to abide by the study protocol, could avoid dropouts.

Panelists also emphasized that methodological issues that have not been completely addressed in on-site trials can be magnified in digital trials (Charvet et al., 2015; Fried et al., 2021). For instance, if the process of randomization - allocation concealment - is hacked from the server and publicly exposed, the entire trial can be lost (or, at least, suspended until a new list is generated). Moreover, blinding and sham stimulation issues are not completely resolved issues in on-site trials (Fonteneau et al., 2019), and biases arising from these steps are more likely to occur (e.g., sham stimulation can be unconcealed by measuring the voltage between electrodes (Woods et al., 2016)) and harm the entire study (for example, by exposing methodological vulnerabilities on the Web). Additionally, issues that would be minor in on-site trials might be more relevant in online trials. For instance, stimulation sessions can be performed in on-site trials appropriately and guarantee adherence (Woods et al., 2016), but, in digital trials, some degree of remote monitoring would be necessary for ensuring these aspects.

Finally, there are unique new challenges for digitizing neuromodulation. Even though cyber-hacking is not usually discussed in the environment of clinical trials, government and big company systems are being increasingly hacked. Data anonymity and confidentiality are additional aspects of vulnerability more relevant than in on-site trials, if, for instance, information is also recorded in the devices, smartphones, or is transmitted remotely. Data collection using standard behavioral scales (for instance, scales for depression) and adverse events need to be further validated to be used online and remotely to avoid instrumental biases. Finally, even open pre-publication of study protocols, which enhances transparency and reproducibility, cannot be fully detailed in digital trials, as a complete description of the groups, blinding methods, and sham stimulation of the protocols could be harmful to the internal validity of digital trials (Charvet et al., 2020, 2015).

### 4.3. Limitations

Although we systematically reviewed the literature for selecting the most productive authors in neuromodulation, experts publishing in non-English databases might not have been selected. In addition, most of the panelists are from high-income countries, limiting the experience, feedback, and the number of votes of panelists from low-/middle-income countries, where 85% of the world population lives, and with probably additional issues on digitalization, including availability. Moreover, our search might not have identified emerging young experts as we have established a threshold based on the number of publications. Although we considered different methods for composing the EP, such as “snowball sampling” based on recommendations from the SC members, and search of other databases such as clinicaltrials.gov, preprints, and conference publications, these processes would be non-systematic or involve gray areas in the literature. We also did not assess other stakeholders besides people from industry and academia that could have been relevant for our work, such as patients, governmental and non-governmental organizations. Moreover, only 8 of 13 companies replied to our contacts, despite several emails that were sent to reach them, and even offering the possibility of online meetings to discuss this work. Although we could have extracted company information based on public information, we opted not to do that as some information could have been inaccurate. Finally, no large tES digital trials have been finished and published yet; therefore, the processes and challenges described here are mostly theoretical and should be iteratively updated as the field develops. Interestingly, the lack of consensus on issues related to its disadvantages, risks, and biases, with many specialists remaining undecided, indicates that the field is still in its infancy.

### 4.4. Future directions

The recommendations for teams conducting digital neuromodulation trials markedly diverge from on-site trials that are centered on clinical specialists and staff trained in performing biomedical procedures. The feasibility of these recommendations should be further debated, as they would require more resources. Most recommendations fit with companies and for pivotal studies, and not necessarily for teams running pilot studies that would not have all the capabilities recommended above. For instance, third-party services could be contracted to do specific tasks related to software and hardware development, or such aspects could be developed together by researchers and companies. Moreover, recommendations such as a support team being always available for medical urgencies, although optimal, might be unrealistic even with large resources. Such issues would need to be carefully discussed with internal review boards and ethics committees to guarantee patient safety without harming trial feasibility.

The recommendations for further research in some aspects specifically related to internal validity, and also external validity, were largely convergent, reaching 70-80% agreement rates. Taken together, these recommendations call for new standards and best practices of fundamental pillars of tES clinical research, such as methods for sham stimulation, randomization, allocation, and assessment of adverse events. These methodologies have been steadily built over the last decade (Bikson et al., 2018a; Brunoni et al., 2019, 2012) and, although challenged in certain aspects such as sham and blinding (Fonteneau et al., 2019; Turner et al., 2021), they have been largely used in clinical trials (Fried et al., 2021). Although several approaches could be used, in a first step relatively simple randomized studies involving healthy participants could use parameters such as blinding efficacy and rate of adverse events as outcome measures, comparing whether they are different in those receiving on-site vs. online tES. Pilot studies using mobile tES in clinical samples are also encouraged to report their methodological approaches and challenges (Alonzo et al., 2019; Eilam-Stock et al., 2021).

## 5. Conclusion

In this first Delphi Panel evaluating opportunities, risks, and methodological issues regarding digitizing tES trials, we provided a landscape of this new approach and reached a consensus on several recommendations that should be evaluated in further studies. The panel of specialists agreed on the advantages associated with the implementation of tES trials; however, considering the fast-growing digitalization in Medicine and Biotechnology, there is a pressing need to better understand how to adapt tES trials to be performed remotely, with a clearer knowledge regarding its positive and negative aspects.

## Data Availability

All data produced in the present study are available upon reasonable request to the authors

## References

ALHarbi, M.F., Armijo-Olivo, S., Kim, E.S., 2017. Transcranial direct current stimulation (tDCS) to improve naming ability in post-stroke aphasia: A critical review. Behav Brain Res 332, 7–15. https://doi.org/10.1016/j.bbr.2017.05.050

Aljedaani, B., Babar, M.A., 2021. Challenges With Developing Secure Mobile Health Applications: Systematic Review. JMIR Mhealth Uhealth 9, e15654. https://doi.org/10.2196/15654

Alonzo, A., Fong, J., Ball, N., Martin, D., Chand, N., Loo, C., 2019. Pilot trial of home-administered transcranial direct current stimulation for the treatment of depression. J Affect Disord 252, 475–483. https://doi.org/10.1016/j.jad.2019.04.041

Antal, A., Alekseichuk, I., Bikson, M., Brockmöller, J., Brunoni, A.R., Chen, R., Cohen, L.G., Dowthwaite, G., Ellrich, J., Flöel, A., Fregni, F., George, M.S., Hamilton, R., Haueisen, J., Herrmann, C.S., Hummel, F.C., Lefaucheur, J.P., Liebetanz, D., Loo, C.K., McCaig, C.D., Miniussi, C., Miranda, P.C., Moliadze, V., Nitsche, M.A., Nowak, R., Padberg, F., Pascual-Leone, A., Poppendieck, W., Priori, A., Rossi, S., Rossini, P.M., Rothwell, J., Rueger, M.A., Ruffini, G., Schellhorn, K., Siebner, H.R., Ugawa, Y., Wexler, A., Ziemann, U., Hallett, M., Paulus, W., 2017. Low intensity transcranial electric stimulation: Safety, ethical, legal regulatory and application guidelines. Clin Neurophysiol 128, 1774–1809. https://doi.org/10.1016/j.clinph.2017.06.001

Antich-Isern, P., Caro-Barri, J., Aparicio-Blanco, J., 2021. The combination of medical devices and medicinal products revisited from the new European legal framework. Int J Pharm 607, 120992. https://doi.org/10.1016/j.ijpharm.2021.120992

Aparício, L.V.M., Guarienti, F., Razza, L.B., Carvalho, A.F., Fregni, F., Brunoni, A.R., 2016. A Systematic Review on the Acceptability and Tolerability of Transcranial Direct Current Stimulation Treatment in Neuropsychiatry Trials. Brain Stimul 9, 671–681. https://doi.org/10.1016/j.brs.2016.05.004

Baptista, A.F., Fernandes, A.M.B.L., Sá, K.N., Okano, A.H., Brunoni, A.R., Lara-Solares, A., Jreige Iskandar, A., Guerrero, C., Amescua-García, C., Kraychete, D.C., Caparelli-Daquer, E., Atencio, E., Piedimonte, F., Colimon, F., Hazime, F.A., Garcia, J.B.S., Hernández-Castro, J.J., Cantisani, J.A.F., Karina do Monte-Silva, K., Lemos Correia, L.C., Gallegos, M.S., Marcolin, M.A., Ricco, M.A., Cook, M.B., Bonilla, P., Schestatsky, P., Galhardoni, R., Silva, V., Delgado Barrera, W., Caumo, W., Bouhassira, D., Chipchase, L.S., Lefaucheur, J.-P., Teixeira, M.J., de Andrade, D.C., 2019. Latin American and Caribbean consensus on noninvasive central nervous system neuromodulation for chronic pain management (LAC2-NIN-CP). Pain Rep 4, e692. https://doi.org/10.1097/PR9.0000000000000692

Bikson, M., Brunoni, A.R., Charvet, L.E., Clark, V.P., Cohen, L.G., Deng, Z.-D., Dmochowski, J., Edwards, D.J., Frohlich, F., Kappenman, E.S., Lim, K.O., Loo, C., Mantovani, A., McMullen, D.P., Parra, L.C., Pearson, M., Richardson, J.D., Rumsey, J.M., Sehatpour, P., Sommers, D., Unal, G., Wassermann, E.M., Woods, A.J., Lisanby, S.H., 2018a. Rigor and reproducibility in research with transcranial electrical stimulation: An NIMH-sponsored workshop. Brain Stimulation: Basic, Translational, and Clinical Research in Neuromodulation 11, 465–480. https://doi.org/10.1016/j.brs.2017.12.008

Bikson, M., Grossman, P., Thomas, C., Zannou, A.L., Jiang, J., Adnan, T., Mourdoukoutas, A.P., Kronberg, G., Truong, D., Boggio, P., Brunoni, A.R., Charvet, L., Fregni, F., Fritsch, B., Gillick, B., Hamilton, R.H., Hampstead, B.M., Jankord, R., Kirton, A., Knotkova, H., Liebetanz, D., Liu, A., Loo, C., Nitsche, M.A., Reis, J., Richardson, J.D., Rotenberg, A., Turkeltaub, P.E., Woods, A.J., 2016. Safety of Transcranial Direct Current Stimulation: Evidence Based Update 2016. Brain Stimul 9, 641–661. https://doi.org/10.1016/j.brs.2016.06.004

Bikson, M., Hanlon, C.A., Woods, A.J., Gillick, B.T., Charvet, L., Lamm, C., Madeo, G., Holczer, A., Almeida, J., Antal, A., Ay, M.R., Baeken, C., Blumberger, D.M., Campanella, S., Camprodon, J.A., Christiansen, L., Loo, C., Crinion, J.T., Fitzgerald, P., Gallimberti, L., Ghobadi-Azbari, P., Ghodratitoostani, I., Grabner, R.H., Hartwigsen, G., Hirata, A., Kirton, A., Knotkova, H., Krupitsky, E., Marangolo, P., Nakamura-Palacios, E.M., Potok, W., Praharaj, S.K., Ruff, C.C., Schlaug, G., Siebner, H.R., Stagg, C.J., Thielscher, A., Wenderoth, N., Yuan, T.-F., Zhang, X., Ekhtiari, H., 2020. Guidelines for TMS/tES clinical services and research through the COVID-19 pandemic. Brain Stimul 13, 1124–1149. https://doi.org/10.1016/j.brs.2020.05.010

Bikson, M., Paneri, B., Mourdoukoutas, A., Esmaeilpour, Z., Badran, B.W., Azzam, R., Adair, D., Datta, A., Fang, X.H., Wingeier, B., Chao, D., Alonso-Alonso, M., Lee, K., Knotkova, H., Woods, A.J., Hagedorn, D., Jeffery, D., Giordano, J., Tyler, W.J., 2018b. Limited output transcranial electrical stimulation (LOTES-2017): Engineering principles, regulatory statutes, and industry standards for wellness, over-the-counter, or prescription devices with low risk. Brain Stimulation: Basic, Translational, and Clinical Research in Neuromodulation 11, 134–157. https://doi.org/10.1016/j.brs.2017.10.012

Brunoni, A.R., Nitsche, M.A., Bolognini, N., Bikson, M., Wagner, T., Merabet, L., Edwards, D.J., Valero-Cabre, A., Rotenberg, A., Pascual-Leone, A., Ferrucci, R., Priori, A., Boggio, P.S., Fregni, F., 2012. Clinical research with transcranial direct current stimulation (tDCS): Challenges and future directions. Brain Stimulation: Basic, Translational, and Clinical Research in Neuromodulation 5, 175–195. https://doi.org/10.1016/j.brs.2011.03.002

Brunoni, A.R., Sampaio-Junior, B., Moffa, A.H., Aparício, L.V., Gordon, P., Klein, I., Rios, R.M., Razza, L.B., Loo, C., Padberg, F., Valiengo, L., 2019. Noninvasive brain stimulation in psychiatric disorders: a primer. Braz J Psychiatry 41, 70–81. https://doi.org/10.1590/1516-4446-2017-0018

Buch, E.R., Santarnecchi, E., Antal, A., Born, J., Celnik, P.A., Classen, J., Gerloff, C., Hallett, M., Hummel, F.C., Nitsche, M.A., Pascual-Leone, A., Paulus, W.J., Reis, J., Robertson, E.M., Rothwell, J.C., Sandrini, M., Schambra, H.M., Wassermann, E.M., Ziemann, U., Cohen, L.G., 2017. Effects of tDCS on motor learning and memory formation: A consensus and critical position paper. Clin Neurophysiol 128, 589–603. https://doi.org/10.1016/j.clinph.2017.01.004

Cappon, D., Jahanshahi, M., Bisiacchi, P., 2016. Value and Efficacy of Transcranial Direct Current Stimulation in the Cognitive Rehabilitation: A Critical Review Since 2000. Front Neurosci 10, 157. https://doi.org/10.3389/fnins.2016.00157

Charvet, L.E., Kasschau, M., Datta, A., Knotkova, H., Stevens, M.C., Alonzo, A., Loo, C., Krull, K.R., Bikson, M., 2015. Remotely-supervised transcranial direct current stimulation (tDCS) for clinical trials: guidelines for technology and protocols. Frontiers in Systems Neuroscience 9.

Charvet, L.E., Shaw, M.T., Bikson, M., Woods, A.J., Knotkova, H., 2020. Supervised transcranial direct current stimulation (tDCS) at home: A guide for clinical research and practice. Brain Stimulation: Basic, Translational, and Clinical Research in Neuromodulation 13, 686–693. https://doi.org/10.1016/j.brs.2020.02.011

Cruccu, G., Garcia-Larrea, L., Hansson, P., Keindl, M., Lefaucheur, J.-P., Paulus, W., Taylor, R., Tronnier, V., Truini, A., Attal, N., 2016. EAN guidelines on central neurostimulation therapy in chronic pain conditions. Eur J Neurol 23, 1489–1499. https://doi.org/10.1111/ene.13103

Darrow, J.J., Avorn, J., Kesselheim, A.S., 2021. FDA Regulation and Approval of Medical Devices: 1976-2020. JAMA 326, 420–432. https://doi.org/10.1001/jama.2021.11171

Deer, T.R., Mekhail, N., Provenzano, D., Pope, J., Krames, E., Leong, M., Levy, R.M., Abejon, D., Buchser, E., Burton, A., Buvanendran, A., Candido, K., Caraway, D., Cousins, M., DeJongste, M., Diwan, S., Eldabe, S., Gatzinsky, K., Foreman, R.D., Hayek, S., Kim, P., Kinfe, T., Kloth, D., Kumar, K., Rizvi, S., Lad, S.P., Liem, L., Linderoth, B., Mackey, S., McDowell, G., McRoberts, P., Poree, L., Prager, J., Raso, L., Rauck, R., Russo, M., Simpson, B., Slavin, K., Staats, P., Stanton-Hicks, M., Verrills, P., Wellington, J., Williams, K., North, R., 2014. The Appropriate Use of Neurostimulation of the Spinal Cord and Peripheral Nervous System for the Treatment of Chronic Pain and Ischemic Diseases: The Neuromodulation Appropriateness Consensus Committee. Neuromodulation 17, 515–550. https://doi.org/10.1111/ner.12208

Eilam-Stock, T., George, A., Lustberg, M., Wolintz, R., Krupp, L.B., Charvet, L.E., 2021. Telehealth transcranial direct current stimulation for recovery from Post-Acute Sequelae of SARS-CoV-2 (PASC). Brain Stimul 14, 1520–1522. https://doi.org/10.1016/j.brs.2021.10.381

Ekhtiari, H., Ghobadi-Azbari, P., Thielscher, A., Antal, A., Li, L.M., Shereen, A.D., Cabral-Calderin, Y., Keeser, D., Bergmann, T.O., Jamil, A., Violante, I.R., Almeida, J., Meinzer, M., Siebner, H.R., Woods, A.J., Stagg, C.J., Abend, R., Antonenko, D., Auer, T., Bächinger, M., Baeken, C., Barron, H.C., Chase, H.W., Crinion, J., Datta, A., Davis, M.H., Ebrahimi, M., Esmaeilpour, Z., Falcone, B., Fiori, V., Ghodratitoostani, I., Gilam, G., Grabner, R.H., Greenspan, J.D., Groen, G., Hartwigsen, G., Hauser, T.U., Herrmann, C.S., Juan, C.-H., Krekelberg, B., Lefebvre, S., Liew, S.-L., Madsen, K.H., Mahdavifar-Khayati, R., Malmir, N., Marangolo, P., Martin, A.K., Meeker, T.J., Ardabili, H.M., Moisa, M., Momi, D., Mulyana, B., Opitz, A., Orlov, N., Ragert, P., Ruff, C.C., Ruffini, G., Ruttorf, M., Sangchooli, A., Schellhorn, K., Schlaug, G., Sehm, B., Soleimani, G., Tavakoli, H., Thompson, B., Timmann, D., Tsuchiyagaito, A., Ulrich, M., Vosskuhl, J., Weinrich, C.A., Zare-Bidoky, M., Zhang, X., Zoefel, B., Nitsche, M.A., Bikson, M., 2022a. A checklist for assessing the methodological quality of concurrent tES-fMRI studies (ContES checklist): a consensus study and statement. Nat Protoc 17, 596–617. https://doi.org/10.1038/s41596-021-00664-5

Ekhtiari, H., Tavakoli, H., Addolorato, G., Baeken, C., Bonci, A., Campanella, S., Castelo-Branco, L., Challet-Bouju, G., Clark, V.P., Claus, E., Dannon, P.N., Del Felice, A., den Uyl, T., Diana, M., di Giannantonio, M., Fedota, J.R., Fitzgerald, P., Gallimberti, L., Grall-Bronnec, M., Herremans, S.C., Herrmann, M.J., Jamil, A., Khedr, E., Kouimtsidis, C., Kozak, K., Krupitsky, E., Lamm, C., Lechner, W.V., Madeo, G., Malmir, N., Martinotti, G., McDonald, W.M., Montemitro, C., Nakamura-Palacios, E.M., Nasehi, M., Noël, X., Nosratabadi, M., Paulus, M., Pettorruso, M., Pradhan, B., Praharaj, S.K., Rafferty, H., Sahlem, G., Salmeron, B. jo, Sauvaget, A., Schluter, R.S., Sergiou, C., Shahbabaie, A., Sheffer, C., Spagnolo, P.A., Steele, V.R., Yuan, T., van Dongen, J.D.M., Van Waes, V., Venkatasubramanian, G., Verdejo-García, A., Verveer, I., Welsh, J.W., Wesley, M.J., Witkiewitz, K., Yavari, F., Zarrindast, M.-R., Zawertailo, L., Zhang, X., Cha, Y.-H., George, T.P., Frohlich, F., Goudriaan, A.E., Fecteau, S., Daughters, S.B., Stein, E.A., Fregni, F., Nitsche, M.A., Zangen, A., Bikson, M., Hanlon, C.A., 2019. Transcranial electrical and magnetic stimulation (tES and TMS) for addiction medicine: A consensus paper on the present state of the science and the road ahead. Neuroscience & Biobehavioral Reviews 104, 118–140. https://doi.org/10.1016/j.neubiorev.2019.06.007

Ekhtiari, H., Zare-Bidoky, M., Sangchooli, A., Janes, A.C., Kaufman, M.J., Oliver, J.A., Prisciandaro, J.J., Wüstenberg, T., Anton, R.F., Bach, P., Baldacchino, A., Beck, A., Bjork, J.M., Brewer, J., Childress, A.R., Claus, E.D., Courtney, K.E., Ebrahimi, M., Filbey, F.M., Ghahremani, D.G., Azbari, P.G., Goldstein, R.Z., Goudriaan, A.E., Grodin, E.N., Hamilton, J.P., Hanlon, C.A., Hassani-Abharian, P., Heinz, A., Joseph, J.E., Kiefer, F., Zonoozi, A.K., Kober, H., Kuplicki, R., Li, Q., London, E.D., McClernon, J., Noori, H.R., Owens, M.M., Paulus, M.P., Perini, I., Potenza, M., Potvin, S., Ray, L., Schacht, J.P., Seo, D., Sinha, R., Smolka, M.N., Spanagel, R., Steele, V.R., Stein, E.A., Steins-Loeber, S., Tapert, S.F., Verdejo-Garcia, A., Vollstädt-Klein, S., Wetherill, R.R., Wilson, S.J., Witkiewitz, K., Yuan, K., Zhang, X., Zilverstand, A., 2022b. A methodological checklist for fMRI drug cue reactivity studies: development and expert consensus. Nat Protoc 17, 567–595. https://doi.org/10.1038/s41596-021-00649-4

Fonteneau, C., Mondino, M., Arns, M., Baeken, C., Bikson, M., Brunoni, A.R., Burke, M.J., Neuvonen, T., Padberg, F., Pascual-Leone, A., Poulet, E., Ruffini, G., Santarnecchi, E., Sauvaget, A., Schellhorn, K., Suaud-Chagny, M.-F., Palm, U., Brunelin, J., 2019. Sham tDCS: A hidden source of variability? Reflections for further blinded, controlled trials. Brain Stimul 12, 668–673. https://doi.org/10.1016/j.brs.2018.12.977

Fregni, F., El-Hagrassy, M.M., Pacheco-Barrios, K., Carvalho, S., Leite, J., Simis, M., Brunelin, J., Nakamura-Palacios, E.M., Marangolo, P., Venkatasubramanian, G., San-Juan, D., Caumo, W., Bikson, M., Brunoni, A.R., Neuromodulation Center Working Group, 2021. Evidence-Based Guidelines and Secondary Meta-Analysis for the Use of Transcranial Direct Current Stimulation in Neurological and Psychiatric Disorders. Int J Neuropsychopharmacol 24, 256–313. https://doi.org/10.1093/ijnp/pyaa051

Fregni, F., Nitsche, M., Loo, C.K., Brunoni, A., Marangolo, P., Leite, J., Carvalho, S., Bolognini, N., Caumo, W., Paik, N., Simis, M., Ueda, K., Ekhitari, H., Luu, P., Tucker, D., Tyler, W., Brunelin, J., Datta, A., Juan, C., Venkatasubramanian, G., Boggio, P., Bikson, M., 2015. Regulatory Considerations for the Clinical and Research Use of Transcranial Direct Current Stimulation (tDCS): review and recommendations from an expert panel. Clin Res Regul Aff 32, 22–35. https://doi.org/10.3109/10601333.2015.980944

Fried, P.J., Santarnecchi, E., Antal, A., Bartres-Faz, D., Bestmann, S., Carpenter, L.L., Celnik, P., Edwards, D., Farzan, F., Fecteau, S., George, M.S., He, B., Kim, Y.-H., Leocani, L., Lisanby, S.H., Loo, C., Luber, B., Nitsche, M.A., Paulus, W., Rossi, S., Rossini, P.M., Rothwell, J., Sack, A.T., Thut, G., Ugawa, Y., Ziemann, U., Hallett, M., Pascual-Leone, A., 2021. Training in the practice of noninvasive brain stimulation: Recommendations from an IFCN committee. Clinical Neurophysiology 132, 819–837. https://doi.org/10.1016/j.clinph.2020.11.018

Gillick, B.T., Gordon, A.M., Feyma, T., Krach, L.E., Carmel, J., Rich, T.L., Bleyenheuft, Y., Friel, K., 2018. Non-Invasive Brain Stimulation in Children With Unilateral Cerebral Palsy: A Protocol and Risk Mitigation Guide. Front Pediatr 6, 56. https://doi.org/10.3389/fped.2018.00056

Godeiro, C., França, C., Carra, R.B., Saba, F., Saba, R., Maia, D., Brandão, P., Allam, N., Rieder, C.R.M., Freitas, F.C., Capato, T., Spitz, M., Faria, D.D. de, Cordellini, M., Veiga, B.A.A.G., Rocha, M.S.G., Maciel, R., Melo, L.B.D., Möller, P.D.S., R R Júnior, M., Fornari, L.H.T., Mantese, C.E., Barbosa, E.R., Munhoz, R.P., Coletta, M.V.D., Cury, R.G., 2021. Use of non-invasive stimulation in movement disorders: a critical review. Arq Neuropsiquiatr 79, 630–646. https://doi.org/10.1590/0004-282X-ANP-2020-0381

Grimaldi, G., Argyropoulos, G.P., Boehringer, A., Celnik, P., Edwards, M.J., Ferrucci, R., Galea, J.M., Groiss, S.J., Hiraoka, K., Kassavetis, P., Lesage, E., Manto, M., Miall, R.C., Priori, A., Sadnicka, A., Ugawa, Y., Ziemann, U., 2014. Non-invasive cerebellar stimulation--a consensus paper. Cerebellum 13, 121–138. https://doi.org/10.1007/s12311-013-0514-7

Hsu, C.-C., Sandford, B., 2019. The Delphi Technique: Making Sense of Consensus. Practical Assessment, Research, and Evaluation 12. https://doi.org/10.7275/pdz9-th90

Inan, O.T., Tenaerts, P., Prindiville, S.A., Reynolds, H.R., Dizon, D.S., Cooper-Arnold, K., Turakhia, M., Pletcher, M.J., Preston, K.L., Krumholz, H.M., Marlin, B.M., Mandl, K.D., Klasnja, P., Spring, B., Iturriaga, E., Campo, R., Desvigne-Nickens, P., Rosenberg, Y., Steinhubl, S.R., Califf, R.M., 2020. Digitizing clinical trials. npj Digit. Med. 3, 1–7. https://doi.org/10.1038/s41746-020-0302-y

Insel, T.R., 2018. Digital phenotyping: a global tool for psychiatry. World Psychiatry 17, 276–277. https://doi.org/10.1002/wps.20550

Kim, D.-J., Moussa-Tooks, A.B., Bolbecker, A.R., Apthorp, D., Newman, S.D., O’Donnell, B.F., Hetrick, W.P., 2020. Cerebellar-cortical dysconnectivity in resting-state associated with sensorimotor tasks in schizophrenia. Hum Brain Mapp 41, 3119–3132. https://doi.org/10.1002/hbm.25002

Lefaucheur, J.-P., Antal, A., Ayache, S.S., Benninger, D.H., Brunelin, J., Cogiamanian, F., Cotelli, M., De Ridder, D., Ferrucci, R., Langguth, B., Marangolo, P., Mylius, V., Nitsche, M.A., Padberg, F., Palm, U., Poulet, E., Priori, A., Rossi, S., Schecklmann, M., Vanneste, S., Ziemann, U., Garcia-Larrea, L., Paulus, W., 2017. Evidence-based guidelines on the therapeutic use of transcranial direct current stimulation (tDCS). Clin Neurophysiol 128, 56–92. https://doi.org/10.1016/j.clinph.2016.10.087

Legatt, A.D., Emerson, R.G., Epstein, C.M., MacDonald, D.B., Deletis, V., Bravo, R.J., López, J.R., 2016. ACNS Guideline: Transcranial Electrical Stimulation Motor Evoked Potential Monitoring. J Clin Neurophysiol 33, 42–50. https://doi.org/10.1097/WNP.0000000000000253

Levac, D., Colquhoun, H., O’Brien, K.K., 2010. Scoping studies: advancing the methodology. Implement Sci 5, 69. https://doi.org/10.1186/1748-5908-5-69

Lucchiari, C., Sala, P.M., Vanutelli, M.E., 2018. Promoting Creativity Through Transcranial Direct Current Stimulation (tDCS). A Critical Review. Front Behav Neurosci 12, 167. https://doi.org/10.3389/fnbeh.2018.00167

Maatoug, R., Bihan, K., Duriez, P., Podevin, P., Silveira-Reis-Brito, L., Benyamina, A., Valero-Cabré, A., Millet, B., 2021. Non-invasive and invasive brain stimulation in alcohol use disorders: A critical review of selected human evidence and methodological considerations to guide future research. Comprehensive Psychiatry 109, 152257. https://doi.org/10.1016/j.comppsych.2021.152257

Martelletti, P., Jensen, R.H., Antal, A., Arcioni, R., Brighina, F., de Tommaso, M., Franzini, A., Fontaine, D., Heiland, M., Jürgens, T.P., Leone, M., Magis, D., Paemeleire, K., Palmisani, S., Paulus, W., May, A., European Headache Federation, 2013. Neuromodulation of chronic headaches: position statement from the European Headache Federation. J Headache Pain 14, 86. https://doi.org/10.1186/1129-2377-14-86

McClintock, S.M., Kallioniemi, E., Martin, D.M., Kim, J.U., Weisenbach, S.L., Abbott, C.C., 2019. A Critical Review and Synthesis of Clinical and Neurocognitive Effects of Noninvasive Neuromodulation Antidepressant Therapies. Focus (Am Psychiatr Publ) 17, 18–29. https://doi.org/10.1176/appi.focus.20180031

Moffa, A.H., Brunoni, A.R., Fregni, F., Palm, U., Padberg, F., Blumberger, D.M., Daskalakis, Z.J., Bennabi, D., Haffen, E., Alonzo, A., Loo, C.K., 2017. Safety and acceptability of transcranial direct current stimulation for the acute treatment of major depressive episodes: Analysis of individual patient data. J Affect Disord 221, 1–5. https://doi.org/10.1016/j.jad.2017.06.021

Onodera, R., Sengoku, S., 2018. Innovation process of mHealth: An overview of FDA-approved mobile medical applications. Int J Med Inform 118, 65–71. https://doi.org/10.1016/j.ijmedinf.2018.07.004

Parikh, S.V., Quilty, L.C., Ravitz, P., Rosenbluth, M., Pavlova, B., Grigoriadis, S., Velyvis, V., Kennedy, S.H., Lam, R.W., MacQueen, G.M., Milev, R.V., Ravindran, A.V., Uher, R., CANMAT Depression Work Group, 2016. Canadian Network for Mood and Anxiety Treatments (CANMAT) 2016 Clinical Guidelines for the Management of Adults with Major Depressive Disorder: Section 2. Psychological Treatments. Can J Psychiatry 61, 524–539. https://doi.org/10.1177/0706743716659418

Polhemus, A.M., Novák, J., Ferrao, J., Simblett, S., Radaelli, M., Locatelli, P., Matcham, F., Kerz, M., Weyer, J., Burke, P., Huang, V., Dockendorf, M.F., Temesi, G., Wykes, T., Comi, G., Myin-Germeys, I., Folarin, A., Dobson, R., Manyakov, N.V., Narayan, V.A., Hotopf, M., 2020. Human-Centered Design Strategies for Device Selection in mHealth Programs: Development of a Novel Framework and Case Study. JMIR Mhealth Uhealth 8, e16043. https://doi.org/10.2196/16043

Razza, L., Brunoni, A., Suen, P.J.C., Ekhtiari, H., Zare-Bidoky, M., Ghobadi-Azbari, P., Zonoozi, A.K., 2021. Digitizing Portable Transcranial Electric Stimulation clinical trials: Scoping Review, Process Mapping, and recommendations from a Delphi Panel. https://doi.org/10.17605/OSF.IO/K83VP

Sanches, C., Stengel, C., Godard, J., Mertz, J., Teichmann, M., Migliaccio, R., Valero-Cabré, A., 2021. Past, Present, and Future of Non-invasive Brain Stimulation Approaches to Treat Cognitive Impairment in Neurodegenerative Diseases: Time for a Comprehensive Critical Review. Frontiers in Aging Neuroscience 12.

Sandars, M., Cloutman, L., Woollams, A.M., 2016. Taking Sides: An Integrative Review of the Impact of Laterality and Polarity on Efficacy of Therapeutic Transcranial Direct Current Stimulation for Anomia in Chronic Poststroke Aphasia. Neural Plast 2016, 8428256. https://doi.org/10.1155/2016/8428256

Santos, F.H., Mosbacher, J.A., Menghini, D., Rubia, K., Grabner, R.H., Cohen Kadosh, R., 2021. Effects of transcranial stimulation in developmental neurocognitive disorders: A critical appraisal. Prog Brain Res 264, 1–40. https://doi.org/10.1016/bs.pbr.2021.01.012

Shiozawa, P., Cordeiro, Q., Cho, H.J., Trevizol, A.P., Brietzke, E., 2017. A critical review of trials of transcranial direct current stimulation and trigeminal nerve stimulation for depression: the issue of treatment-emergent mania. Trends Psychiatry Psychother 39, 48–53. https://doi.org/10.1590/2237-6089-2016-0027

Sierawska, A., Prehn-Kristensen, A., Moliadze, V., Krauel, K., Nowak, R., Freitag, C.M., Siniatchkin, M., Buyx, A., 2019. Unmet Needs in Children With Attention Deficit Hyperactivity Disorder-Can Transcranial Direct Current Stimulation Fill the Gap? Promises and Ethical Challenges. Front Psychiatry 10, 334. https://doi.org/10.3389/fpsyt.2019.00334

Silva-Filho, E., Pilloni, G., Charvet, L.E., Fregni, F., Brunoni, A.R., Bikson, M., 2022. Factors supporting availability of home-based Neuromodulation using remote supervision in middle-income countries; Brazil experience. Brain Stimulation: Basic, Translational, and Clinical Research in Neuromodulation 15, 385–387. https://doi.org/10.1016/j.brs.2022.02.005

Thibaut, A., O’Brien, A.T., Fregni, F., 2017. Strategies for replacing non-invasive brain stimulation sessions: recommendations for designing neurostimulation clinical trials. Expert Rev Med Devices 14, 633–649. https://doi.org/10.1080/17434440.2017.1352470

Torous, J., Bucci, S., Bell, I.H., Kessing, L.V., Faurholt-Jepsen, M., Whelan, P., Carvalho, A.F., Keshavan, M., Linardon, J., Firth, J., 2021. The growing field of digital psychiatry: current evidence and the future of apps, social media, chatbots, and virtual reality. World Psychiatry 20, 318–335. https://doi.org/10.1002/wps.20883

Torous, J., Onnela, J.-P., Keshavan, M., 2017. New dimensions and new tools to realize the potential of RDoC: digital phenotyping via smartphones and connected devices. Transl Psychiatry 7, e1053. https://doi.org/10.1038/tp.2017.25

Turner, C., Jackson, C., Learmonth, G., 2021. Is the “end-of-study guess” a valid measure of sham blinding during transcranial direct current stimulation? Eur J Neurosci 53, 1592–1604. https://doi.org/10.1111/ejn.15018

Woods, A.J., Antal, A., Bikson, M., Boggio, P.S., Brunoni, A.R., Celnik, P., Cohen, L.G., Fregni, F., Herrmann, C.S., Kappenman, E.S., Knotkova, H., Liebetanz, D., Miniussi, C., Miranda, P.C., Paulus, W., Priori, A., Reato, D., Stagg, C., Wenderoth, N., Nitsche, M.A., 2016. A technical guide to tDCS, and related non-invasive brain stimulation tools. Clinical Neurophysiology 127, 1031–1048. https://doi.org/10.1016/j.clinph.2015.11.012

Workman, C.D., Fietsam, A.C., Rudroff, T., 2020. Tolerability and Blinding of Transcranial Direct Current Stimulation in People with Parkinson’s Disease: A Critical Review. Brain Sci 10, E467. https://doi.org/10.3390/brainsci10070467

Zhang, Y., Song, H., Chen, Y., Zuo, L., Xia, X., Zhang, X., 2019. Thinking on Transcranial Direct Current Stimulation (tDCS) in Reading Interventions: Recommendations for Future Research Directions. Front Hum Neurosci 13, 157. https://doi.org/10.3389/fnhum.2019.00157

