## Supplementary Material for "Digitizing Non-Invasive Neuromodulation Trials: Scoping Review, Process Mapping, and Recommendations from a Delphi Panel" (from another version of this preprint: version 1, 2022-03-05)

### Digitizing Portable Transcranial Electric Stimulation clinical trials: Scoping Review, Process Mapping, and recommendations from a Delphi Panel.

**Appendix 1. Deviation from the pre-registered protocol**

**Appendix 2. Steering Committee and Expert Panel questionnaire**

**Appendix 3. Simulations and Thresholds for selection of the Expert Panel**

**Appendix 4.  Systematic scoping review**

**Appendix 5. tES companies questionnaire**

**Appendix 6. Steering Committee and Expert Panel Working Group**

**Appendix 7. Demographic characteristics of the included authors**

**Appendix 8. Systematic scoping review results**

**Appendix 9. Rating scores for the SWOT assessment**

**Appendix 10. Rating scores for the SIPOC assessments**

**Appendix 11. Rating scores for general recommendations**

#### Appendix 1. Deviation from the pre-registered protocol

##### Scoping Review

For the scoping review we have not included all sources of secondary data as we have previously described in the pre-registered protocol. We have decided to include only recommendation papers, guidelines, critical reviews and consensus papers since they present a more critical overview of the literature. Therefore, meta-analysis, umbrella reviews and literature reviews were not selected.

##### Panel of specialists

For the expert panel selection we included authors with at least 3 papers (as first, last or corresponding author). The pre-registered protocol was to have at least 2 papers, howerever, since the first pre-registered search yielded a high number of experts and the selection of the expert panel was unfeasible for this project, the threshold was changed to at least three papers based on the decisions made by the steering committee.

##### Questionnaires

During the questionnaire rating phase, the Steering Committee and the Expert Panel had to answer each question based on a Five-point-Likert-Scale with the following possibilities: "*Strongly Disagree", "Disagree", "Neutral", "Agree" and "Strongly Agree".* This was slightly different from the pre-registered protocol, in which we have included a Likert scale including answers like *"Not Important", "Slightly Important", "Moderately Important", "Very Important", and "Essential".*

##### Assessment of digitalization readiness of tES devices.

The process describing the assessment of digitalization readiness was absent in the original protocol. It was added during the process of analyzing the results for a better intuition of the overall digitalization aspects of tES. The employed methodology and the original results are presented below.

**Appendix 2. Steering Committee and Expert Panel questionnaire**

In a first step, members of the Steering Committee (SC) discussed several items using semi-structured questionnaires that were prepared by the main authors after reviewing the articles from the scoping review. This interaction was performed using electronic forms in which the SC could comment and suggest exclusion or inclusion of new items. At this step, some items were not included since they did not reach a minimum 60% threshold and others were suggested and further added. Upon completion of this phase, both the EP and SC members rated each item according to a Five-point Likert scale (**Supplementary Table 1**). The SWOT (Strengths, Weaknesses, Opportunities and Threats) and the SIPOC (Suppliers, Inputs, Process, Outputs, Customers) approaches were used in the questionnaire to respectively identify external and internal negative and positive aspects for digitizing tES trials. Afterward, an initial draft version of the manuscript was prepared by the main authors and revised by all co-authors, until a final version was achieved. At this step, additional information regarding commercial tES devices were also inserted and discussed. The SC members were identified using a snowball methodology based on the collaborative network of the main authors.

(1) SWOT (Strengths, Weaknesses, Opportunities and Threats) assessment. The SWOT approach is a business strategy tool to identify issues in a project [1]. "Strengths" and "Weaknesses" are internally-related. Here, they identify inherent characteristics of conventional and portable tES that can either facilitate or impede digital trials. "Opportunities" and "Threats" were used to identify external elements that can be exploited or can be harmful for implementing tES digital trials.

(2) SIPOC (Suppliers, Inputs, Process, Outputs, Customers) assessment. The SIPOC tool from Lean and Six Sigma [2] is used to describe processes, and employed here to compare the main processes involved in standard and digital tES trials.

**Supplementary Table 1. Five-point Likert questionnaire addressed to both Steering Committee and Expert Panel members.**

| |
|---|
| Please note that all the information you provide in this section and all the other ones will be treated as confidential and used only by the coordinators of the study and their team. Only summary data will be analyzed. |
| Your name |
| Your e-mail |
| Where is your lab located? |
| How many years of experience do you have in the NIBS field? |
| What is your gender? |
| What is your age group? |
| What is your education level? |
| Country of residence |
| Primary affiliation |
| Primary field(s) of research (except for NIBS) |
| Primary place(s) of work |
| Experience in conducting tDCS or rTMS clinical trials (years) |
| Experience in conducting tDCS or rTMS clinical trials using digital features (years) |
| Number of clinical trials using tDCS that you have already conducted as a PI or Co-I |
| Number of clinical trials using tDCS that you have already been involved with (as a consultant, external advisor, team member, data analyst, etc) |

| |
|---|
| Number of clinical trials using tDCS and digital features that you are currently conducting as a PI or Co-I |
| Number of clinical trials using tDCS and digital features that you are currently involved with (e.g., as a consultant, external advisor, team member, etc) |
| Are you planning to start a digital trial using ptES in the next 2 years? |
| We invited 6 companies that we identified that are major playes in ptES devices: Soterix, Flow Neuroscience, Neuroconn, YBrain, Neurolectrics, and Sooma. Would you like to suggest another company that we should contact? In this case, it would be helpful to know if you can also help us liaise to them. |
| **Swot Assessment*** <br> ***All items were rated each according to a Five-point Likert scale: Strongly Disagree, Disagree, Neutral, Agree, Strongly Agree** |
| **Strengths and Opportunities** |
| Safety of performing ptES sessions outside of clinical settings |
| Simplicity of self-performing ptES sessions outside of clinical settings |
| tES and ptES being a non-pharmacological intervention (instead of a pharmacological intervention) |
| ptES devices being a non-expensive intervention |
| ptES being a scalable intervention - or having "low marginal costs" - i.e., the costs of enrolling patients relatively decrease (in terms of costs per patient) as the number of patient increases |
| Use of telemedicine/telehealth in ptES digital trials |
| Use of mobile Health techonologies for performing digital phenotyping (i.e., quantifying behaviors using real-time measurements) |
| Use of mobile Health techonologies to combine ptES with digital interventions (such as digital therapies) |
| Automatization of several processes within a clinical trial (such as recruitment, asessments, follow-up, and others) |
| Use of 5G / Internet of Things to allow seamless comunication between ptES devices and other portable, digital technologies |
| Developing more customized devices using design thinking strategies (please check the concept, if you are unfamiliar with it, in https://www.frontiersin.org/articles/10.3389/fpubh.2020.00459/full) |
| If you wish, please add any comment on the strengths / opportunities listed above. |
| **Weaknesses/Threats** |
| Risks related to hacking and poor cyber-security |
| Risks related to poor confidentiality and anonymity of the data |
| Risks related to "do-it-yourself" or and/or recreational ptES use |
| Risks related to insufficient supervision and monitoring of self-applied ptES sessions |
| Bioethical issues (e.g., communities from low-/middle- income countries or with low digital literacy would not have access to ptES) |
| Difficulties in obtaining accurate electrode placement in ptES sessions |
| Skin burns as a particularly relevant side effect of ptES digital trials |
| Unclear regulatory status of ptES / tDCS devices |
| Lack of evidence of efficacy for most clinical conditions |
| Risks related to serious adverse events that would be more likely to happen / more difficult to handle in ptES digital trials (e.g., manic switch or suicidal ideation) |
| If you wish, please add any comment on the aspects listed above. |
| **SIPOC analysis of tDCS / ptES clinical trials*** <br> ***All items were rated each according to a Five-point Likert scale: Strongly Disagree, Disagree, Neutral, Agree, Strongly Agree** |
| The main differences between the recruitment processes of pTES trials in which digital features are absent or present are accurately represented. |
| If you wish, please add comments to the question above. |
| The main differences between the pre-screening processes of pTES trials in which digital features are absent or present are accurately represented. |
| If you wish, please add comments to the question above. |
| The main differences between the screening processes of ptES trials in which digital features are absent or present are accurately represented. |
| If you wish, please add comments to the question above. |
| The main differences between the processes of participation in ptES trials in which digital features are absent or present are accurately represented. |

| |
|---|
| If you wish, please add comments to the question above. |
| Any study design (i.e., parallel, cross-over, factorial etc) can be employed in ptES digital trials. |
| Complex study designs (adaptive clinical trials) can be used in ptES digital trials. |
| ptES digital trials should be performed only in conditions that have being already evaluated in on-site pilot studies (e.g., if one wishes to test a new protocol for tinnitus, initially a on-site trial must be performed) |
| ptES digital trials should employ new methods for assessing clinical effects and adverse effects |
| ptES digital trials present increased risk (compared with on-site tDCS trials) of not detecting suicidal behaviors during the trial |
| ptES digital trials present increased risk (compared with on-site tDCS trials) of not detecting manic switches during the trial |
| ptES digital trials present increased risk (compared with on-site tDCS trials) of not detecting adverse cognitive effects during the trial |
| ptES digital trials present increased risk (compared with on-site tDCS trials) of not detecting serious adverse effects during the trial |
| ptES devices have already an adequate level of development that permit performing ptES digital trials |
| In the next decade, ptES digital trials will fully substitute on-site tDCS trials, making them no longer necessary. |
| ptES digital trials can follow-up participants for longer periods than on-site trials |
| ptES digital trials can recruit more participants compared to on-site trials |
| ptES digital trials are faster to be conducted than on-site trials |
| ptES have greater external validity (generalizability) than on-site trials. Here, consider that, on one hand, on-site trials have limited catchment area and usually enroll people that are able to go to the clinical center (potentially less severe and/or with more free time), barriers that could be surpassed by digital trials; and, on the other hand, that ptES digital trials might recruit samples with higher level of digital literacy (potentially younger and richer) |
| ptES digital trials are likely to present increased attrition rates (more drop-outs) compared to on-site trials. Here, consider that, on one hand, digital trials do not require people to return to the clinical center every day; and that, on the other hand, on-site trials might promote greater engagement due to daily interactions of the clinical team. |
| ptES digital trials have greater internal validity (i.e., less risks of methodological biases) than on-site trials. |
| ptES digital trials are more vulnerable to selection biases (i.e. selecting sample with specific characteristics that might not represent the universe of the condition being studied) than on-site trials |
| ptES digital trials are more vulnerable to randomization/allocation biases (i.e. a set of processes that do not allow researchers and participants to be aware of the group that the next participant will be allocated to) than on-site trials. Here, consider the potential risk of exposing the allocation group using social media, discussing the allocation group in forums such as reddit, and potentially hacking the server of the research center to have access of the full list |
| ptES digital trials are more vulnerable to blinding biases (i.e. a set of processes that do not allow researchers and participants to be aware of the group that the enrolled participants are allocated) than on-site trials. Here, consider the aspects mentioned above, and also that electric devices (e.g., voltmeter) can be used to blinding breaking |
| ptES digital trials are more vulnerable to not performing adequate sham stimulation (i.e., increased risk of detection) than on-site trials. Here, consider the issues mentioned above. |
| ptES digital trials are more vulnerable to outcome assessment biases (i.e., measuring the outcome of the study) than on-site trials |
| ptES digital trials can collect more data than on-site trials, particularly ecological momentary data and behaviors during the day |
| ptES digital trials have greater potential of being combined with other therapies simultaneously (such as digital therapies) than on-site trials |
| ptES digital have greater potential of scalability than on-site trials |
| If you wish, please provide additional comments. |
| **Recommendations***<br>***All items were rated each according to a Five-point Likert scale: Strongly Disagree, Disagree, Neutral, Agree, Strongly Agree** |
| The team performing digital trials using pTES should have at least one person skilled in digital marketing strategies for patient recruitment. |
| The team performing digital trials using pTES should have at least one person skilled in data science and data visualization |
| The team performing digital trials using pTES should have at least one person skilled in data science and data visualization (or being advised by such a person) |
| The team performing digital trials using pTES should have at least one person skilled in design thinking (or being advised by such a person) to develop better front-end features and increase usability |
| The team performing digital trials using ptES should have at least one person skilled in app programming (or being advised by such a person) to develop better back-end features and increase usability |

| |
|---|
| The team performing digital trials using ptES should have at least one person skilled in issues related to data security (or being advised by such a person) |
| The team performing digital trials using ptES should have at least one person skilled in issues related to data security (or being advised by such a person) |
| The team performing digital trials using ptES should have a person available at all times (including weekends) to handle emergencies |
| In the field of ptES digital trials, more studies are warranted for assessing randomization and allocation biases and developing better methods for these procedures. |
| In the field of ptES digital trials, more studies are warranted for assessing biases related to sham and developing better methods for this procedure. |
| In the field of ptES digital trials, more studies are warranted for assessing biases related to attrition and developing better methods for this procedure. |
| In the field of ptES digital trials, more studies are warranted for assessing how to enhance external validity of such trials. |
| In the field of ptES digital trials, more studies are warranted for assessing how to combine TES with simultaneous digital interventions |
| In the field of ptES digital trials, more studies are warranted for assessing how to combine TES with ecological data collection |
| In the field of ptES digital trials, more studies are warranted for assessing whether data are securely stored |
| In the field of ptES digital trials, more studies are warranted for assessing how to decrease common adverse effects (e.g., skin reddening, itching, burning, etc) |
| In the field of ptES digital trials, more studies are warranted for assessing how to handle serious adverse events (e.g., manic switch and others) |
| In the field of ptES digital trials, more studies are warranted for assessing unexpected adverse effects that have not yet been described in on-site trials |
| In the field of ptES digital trials, more studies are warranted for assessing biases related to sham and developing better methods for thi procedure. |
| In the field of ptES digital trials, more studies are warranted for assessing / developing better methods of sham. |
| If you wish, please describe and comment additional recommendations. |
| Please check your potential COIs |
| Please add any other comments below. |

**Appendix 3. Simulations and Thresholds for selection of the Expert Panel**

We initially performed a systematic review in PubMed/Medline from January 1st 2012 to September 6th 2021 using keywords and variants related to tES and randomized clinical trials to identify the EP members. We did not take into account studies published before 2012 as we wished to identify only active researchers in the field. This strategy yielded 1120 articles and 4763 authors. Of those, 458 authors (~ 10% of the total sample) published at least 3 papers. Based on different simulations and thresholds, we identified 46 authors, who had at least 7 papers and contributed as the main (first or last) authors of at least 3 papers, to be part of the EP, which approximately corresponds to 2% of the most productive researchers in the field.

**Supplementary Figure 1. Different thresholds for recruiting the expert panel based on the current literature**. Figures (A) to (F) show how many authors have been chosen based on different thresholds between the aforementioned group. Based on the previous experience in conducting Delphi studies[3], the (D) threshold was chosen, i.e. those authors with at least 7 papers and keeping the authorship position of first or last of at least 3 papers.

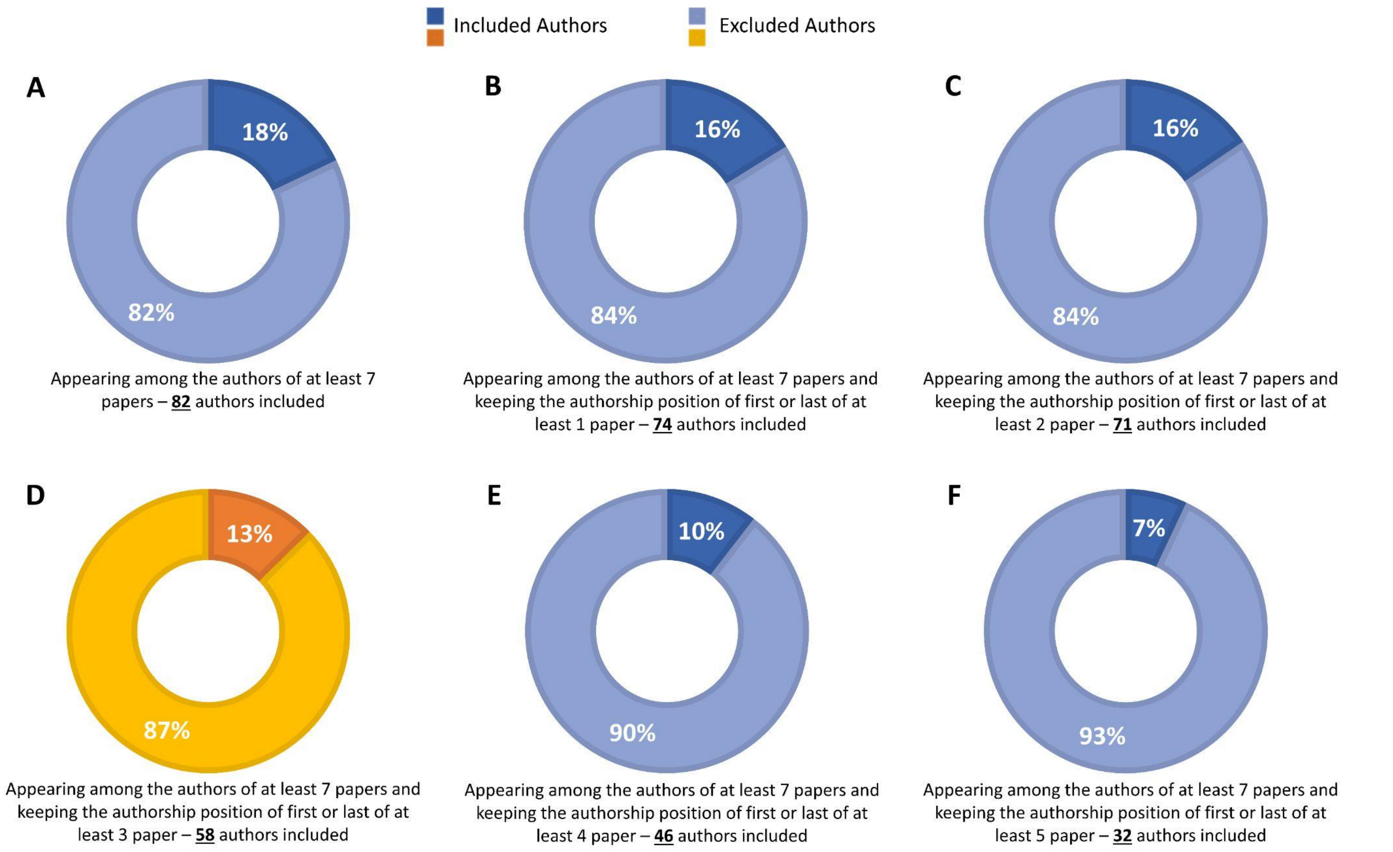

**Appendix 4. Systematic scoping review**

**Search strategy for the systematic reviews**

A systematic scoping review was performed on PubMed/MEDLINE, from January 1st, 2012 to September 9th, 2021, using the keywords “transcranial electric stimulation*”,* “expert consensus”, “guidelines,”, ”recommendations,” and similar terms. This type of review was chosen considering that scoping approaches can provide a broad overview of topics without emphasizing specific study designs, rigid data extraction protocols, or quality assessments while simultaneously retaining other features of a systematic approach on the literature [4]. Therefore, we looked for recent (hence the data range) articles, including large tES trials, methodological systematic reviews, consensus papers to select characteristics to be further evaluated by the first and last authors to compose the questionnaires used in this work.

**Search strategy on PubMED for the scoping review**

('transcranial electric stimulation' OR 'tES' OR 'transcranial direct current stimulation' OR 'tdcs') AND ('consensus' OR 'expert consensus' OR 'guidelines' OR 'guideline' OR 'recommendation' OR 'recommendations' OR 'umbrella review' OR 'critical review')

**Search Strategy on PubMED for the expert panel selection**

((transcranial direct current stimulation) OR (tdcs) OR (transcranial random noise stimulation) OR (trns) OR (transcranial alternating current stimulation) OR (tacs) OR (transcranial electrical stimulation)) AND ((sham-controlled trial) OR (randomized controlled trial) OR (controlled trial) OR (randomized trial))

#### Appendix 5. tES companies questionnaire

We identified companies producing tES devices based on (1) booths of these companies in conferences of the field, (2) references by members of the SC and EP, and (3) Web and Social Media search. The companies were asked regarding general company information, tES montage, characteristics of electrodes and sponges, parameter space of stimulation, safety recommendations, user experience, connectivity, cybersecurity, features for data collection, ecological footprint, and clinical and digital trial features. We do not present data of companies that did not reply to our contacts (initial e-mail contact and two reminders within 2 months).

##### Supplementary Table 2. Questionnaire addressed to the tES companies

| |
|---|
| Timestamp |
| Your Name |
| Name of the Company |
| Name of the (main or more recent one if more than one) portable Transcranial Electric Stimulation (pTES) device |
| Company website |
| Company HD´s country |
| Year the company was founded |
| Has the device received approval from any regulatory agencies? |
| Which regulatory agencies have approved the device? |
| Please upload a pdf with the technical documentation of your device |
| Please upload a picture of the device |
| **Section: Montage** |
| Select the description that best fits the electrode positioning of your device: |
| Please briefly describe the answer above, if necessary |
| Select the best option describing the sponges used by the device:<br>Built-in system<br>Generic straps<br>Customized straps<br>EEG-like headcap<br>Other |
| Please briefly describe the answer above, if necessary |
| Please provide a brief description of the sponge size and material:<br>1-time use (disposable)<br>Re-usable<br>Customized |
| Please provide a brief description of how the sponge is attached to the electrode and how humidification is performed |
| **Section: Parameter space, safety, and usability** |
| Regarding the parameter space: [Dosage] Options: Always fixed, Always adjusted (i.e., is defined for each session), Usually fixed, but can be in some occasions adjusted |
| Regarding the parameter space: [Session duration] Options: Always fixed, Always adjusted (i.e., is defined for each session), Usually fixed, but can be in some occasions adjusted |
| Regarding the parameter space: [Head size] Options: Always fixed, Always adjusted (i.e., is defined for each session), Usually fixed, but can be in some occasions adjusted |
| Regarding the parameter space: [Positioning] Options: Always fixed, Always adjusted (i.e., is defined for each session), Usually fixed, but can be in some occasions adjusted |

| |
|---|
| Regarding the parameter space: [Number of sessions] Options: Always fixed, Always adjusted (i.e., is defined for each session), Usually fixed, but can be in some occasions adjusted |
| Regarding the parameter space: [Combination of therapies] Options: Always fixed, Always adjusted (i.e., is defined for each session), Usually fixed, but can be in some occasions adjusted |
| For the adjustable parameters, please explain how the parameters are programmed and who can program them. |
| Please provide a description of the safety precautions recommended during device use |
| What is the main front-end interface of your device?<br>Smartphone app -4<br>The device itself, Graphical Use Interface, Touch-Screen -3<br>The device itself, no GUI and/or no Touch Screen – 2<br>The device itself, no interactions with the device - 1<br>Other |
| Please provide a description of how the stimulation session starts (e.g., user-dependent, staff-dependent, and how this is done - by app, via a PIN code, etc) |
| Please describe how the session ends |
| What information of the session is displayed to the participant? For instance, is the user warned that the session is starting or ending? If the session is interrupted for some reason, is the user instructed to reconnect the device? |
| How is the battery of the device recharged? |
| Are gamification strategies used to enhance patient´s adherence? (by app, e-mail, messages, etc?) Please describe them. |
| Is data on end-user experience (satisfaction, issues) routinesly collected? |
| Is customer service routinely available? |
| **Section: Connectivity** |
| Does your device operate..<br>100% offline, receives info only via cables, attached modules, or PIN codes (countercoide in the device itself)<br>using Bluetooth<br>using Bluetooth and/or Wi-fi<br>using Bluetooth and/or Wi-fi and/or 3G-4G-LTE<br>Other |
| Does your pTES "talk with" other devices wirelessly?<br>No<br>Yes, with smartphone<br>Yes, with another device from our company<br>Yes, with wearables<br>Other |
| **Section: Data collection and security** |
| Please provide a description of cybersecurity features of the local device, server and cloud data storage (when applicable) |
| Please provide a description of procedures employed to ensure data anonymity and user privacy |
| Is your device / system in accordance with GDPR and/or similar laws? |
| What data is collected by the device? |
| **Section: Ecological footprint** |
| Please provide information regarding device lifetime, battery lifetime and the maximum number of sessions. |
| Can the battery be replaced?<br>Yes<br>No<br>Theoretically, but we have no standardization of this process<br>We have no standardization of this process but we plan to implement it in the near future |
| Can the device be recycled?<br>Yes<br>No<br>Partly |

| |
|---|
| Can the sponges be recycled?<br>Yes<br>No<br>Partly |
| Does your company have specific strategies to reduce its ecological footprint? |
| **Section: Clinical and Digital Trials** |
| Right now, would you say that the interest in conducting on-site clinical trials (together with clinical researchers in the field) by your company is:<br>Very Low<br>Low<br>Neutral<br>High<br>Very High |
| Right now, would you say that the interest in conducting on-line (home-use, remote using portable TES) clinical trials (together with clinical researchers in the field) by your company is:<br>Very Low<br>Low<br>Neutral<br>High<br>Very High |
| What are the main characteristic(s) that would make your company more willing to be involved in any type of clinical trials?<br>Possibility to raise funds (either public or private)<br>Possibility to sponsor our own trials (industry-initiated trials)<br>Possibility to advance our technology collaborating with technical researchers<br>Possibility to advance our technology collaborating with clinicians<br>Possibility to work with leaders of the field<br>Other |
| What features do your devices have and/or can be implemented for randomization and allocation procedures (consider both for on-site and on-line trials)? This procedures describe methods to keep participants and staff unaware of the allocated group of the NEXT participant. |
| What features do your devices have and/or can be implemented for blinding (consider both for on-site and on-line trials)? This procedures describe methods to keep participants and staff unaware of the allocated group of the CURRENT participant. |
| Can your device perform / be adapted to perform sham tDCS / pTES? How is (would be) this performed? |
| What other concomitant interventions, besides tDCS / pTES, can your device do / be adapted to do? |
| What data can be collected by the device in terms of adaptations of software? (consider Chatbots and EMAs)? |
| What data can be collected by the device in terms of adaptations of hardware? (consider EEG and wearables)? |
| What features does your company offer (can offer) for facilitating trial implementation by researchers (consider adaptations of the system in different settings / languages, use of dashboards, data collection and integration with platforms used in clinical research such as R, RedCap, Python, etc) |
| Please describe other aspects that you judge of interest that your device / system/ company can offer in the context of on-site/on-line clinical trials |
| **SWOT assessment**<br>**Strengths** |
| Non-pharmacological device<br>Strongly disagree<br>Disagree<br>Neutral<br>Agree<br>Strongly Agree |
| Safety<br>Strongly disagree<br>Disagree<br>Neutral<br>Agree<br>Strongly Agree |

| |
|---|
| Simplicity (of use)<br>Strongly disagree<br>Disagree<br>Neutral<br>Agree<br>Strongly Agree |
| Non-expensive<br>Strongly disagree<br>Disagree<br>Neutral<br>Agree<br>Strongly Agree |
| Positive phase-III trials<br>Strongly disagree<br>Disagree<br>Neutral<br>Agree<br>Strongly Agree |
| Would you like to add other "strengths" or make additional comments? |
| **Weaknesses** |
| Limited parameter space (for pTES)<br>Strongly disagree<br>Disagree<br>Neutral<br>Agree<br>Strongly Agree |
| Inaccurate (regarding electrode placement)<br>Strongly disagree<br>Disagree<br>Neutral<br>Agree<br>Strongly Agree |
| Skin burns (for pTES)<br>Strongly disagree<br>Disagree<br>Neutral<br>Agree<br>Strongly Agree |
| Regulatory status<br>Strongly disagree<br>Disagree<br>Neutral<br>Agree<br>Strongly Agree |
| Would you like to add other "weaknesses" or make additional comments? |
| **Opportunities** |
| (use of) mobile Health<br>Strongly disagree<br>Disagree<br>Neutral<br>Agree<br>Strongly Agree |

| |
|---|
| Scalability<br>Strongly disagree<br>Disagree<br>Neutral<br>Agree<br>Strongly Agree |
| Telemedicine<br>Strongly disagree<br>Disagree<br>Neutral<br>Agree<br>Strongly Agree |
| Automatization<br>Strongly disagree<br>Disagree<br>Neutral<br>Agree<br>Strongly Agree |
| (use of) 5G/IoT (to leverage use of pTES)<br>Strongly disagree<br>Disagree<br>Neutral<br>Agree<br>Strongly Agree |
| Design thinking (approaches to enhance development)<br>Strongly disagree<br>Disagree<br>Neutral<br>Agree<br>Strongly Agree |
| Would you like to add other "opportunities" or make additional comments? |
| **Threats** |
| Hacking and cyber-security<br>Strongly disagree<br>Disagree<br>Neutral<br>Agree<br>Strongly Agree |
| Data confidentiality and anonimity issues<br>Strongly disagree<br>Disagree<br>Neutral<br>Agree<br>Strongly Agree |
| Disruptions in the supply chain (consider that large-scale pTES trials would be performed with > 500 devices)<br>Strongly disagree<br>Disagree<br>Neutral<br>Agree<br>Strongly Agree |
| Ecological footprint<br>Strongly disagree<br>Disagree<br>Neutral<br>Agree<br>Strongly Agree |
| Would you like to add other "threats" or make additional comments? |

| Please let us know if |
|---|

**Supplementary Table 3.** Overview of available tES devices, as supplied by the companies.

| Name of the Company | Flow Neuroscience | neuroConn GmbH | Ybrain Inc. | Neuroelectrics | Soterix Medical | PlatoScience ApS | EMS srl |
|---|---|---|---|---|---|---|---|
| **Country of origin** | Sweden | Germany | South Korea | Spain | United States | Denmark | Italy |
| **Name of the tES device** | Flow | neuroConn DC-STIMULATOR MOBILE | STIM+ | Starstim-Home tES-EEG Starstim-Home tES | mini-CT | PlatoWork | BrainStim plus |
| **Stimulation parameters for end user** | | | | | | | |
| Electrode positioning | Fixed | Adjusted | Adjusted | Fixed* | Fixed | Adjusted | Adjusted |
| Session duration | Fixed | Adjusted | Adjusted | Fixed* | Adjusted | Adjusted | Adjusted |
| Current intensity | Fixed | Adjusted | Adjusted | Fixed* | Adjusted | Adjusted | Adjusted |
| Combination of therapies | Fixed | Adjusted | Adjusted | Fixed* | Adjusted | Adjusted | Adjusted |
| **Connectivity** | | | | | | | |
| USB | Absent | Present | Absent | Absent | Absent | Absent | Absent |
| Bluetooth | Present | Absent | Present | Absent | Absent | Present | Present |
| Wi-Fi | Absent | Absent | Present | Present | Present | Absent | Absent |
| **User experience** | | | | | | | |
| Stimulation start | User | User | Automatic | User† | User | Automatic | User |
| Stimulation end | Automatic | Automatic | Automatic | Automatic | Automatic | Automatic | Automatic |
| Moisturize sponge before use | Automatic | User | User | User | Automatic | User | User |
| **Cybersecurity features** | | | | | | | |
| User gives consent to collection of their data | Present | Present | Present | Optional | Present | Present | Absent |
| Data is transferred to a cloud server | Present | Present | Present | Present | Present | Present | Absent |
| Data is manually transferred by USB or Bluetooth | Absent | Present | Absent | Absent | Absent | Absent | Present |
| **Data collected by device or app** | | | | | | | |
| Stimulation data (impedance, duration, date, time) | Yes | Yes | Yes | Yes | Yes | Yes | Yes |
| Response to questionnaires | Yes | No | No | Yes | Yes | Yes | No |
| User tolerability and side effects | Yes | No | No | Yes | Yes | Yes | No |
| **Safety recommendations** | | | | | | | |
| Instructs correct positioning of device | Present | Present | Present | Present | Present | Present | Absent |
| Importance of skin cleanness | Present | Absent | Present | Present | Present | Present | Present |
| General contraindications for tES | Present | Present | Present | Present | Present | Present | Present |
| **Ecological footprint** | | | | | | | |
| Sponges can be reutilized after one session | No | Yes | Yes | Yes | No | Yes | Yes |
| Devices can be recycled | Yes | Yes | No | Partly | Partly | Partly | Partly |
| Sponges can be recycled | No | Yes | No | No | No | Yes | No |

| Clinical and digital trial features | | | | | | | |
|---|---|---|---|---|---|---|---|
| Sham stimulation | Present | Present | Present | Present | Present | Present | Present |
| Web-based dashboard management | Present | Absent | Present | Present | Present | Present | Absent |
| Deliver online questionnaires | Present | Absent | Present | Present | Present | Present | Absent |
| **Approval of regulatory agencies** | Yes | Yes | Yes | No | Yes | Yes | Yes |

*can be adjusted for the study by the staff or even individualized based on personalized montages, but are to be delivered to the patient as fixed.

✝importantly, in the time boundaries controlled by the staff.

Fixed/Adjusted: Stimulation parameters can be fixed, or adjusted by the end-user or the company (with an online connection or preset options) after the patient is in possession of the device.

User/Automatic: Procedural steps to prepare and start the stimulation session, either done by the user himself or automatically.

##### tES digitalization readiness

According to the feedback of the companies, we considered the following parameters for measuring digital readiness?

a) Connectivity - we graded one point for each characteristic: presence of Bluetooth, presence of Wi-Fi, presence of 3/4/5G, connectivity to third-party apps (total of 4 points)
b) Readiness for digital trials - if the device had scalable and potentially automatization for the following processes, even if optional (ie, purchased or acquired separately, and not necessarily part of the usual device), it scored one point for each: randomization procedures, sham procedures, blinding of staff, blinding of participants, built-in data collection, biological data collection, and dashboard for clinical trials (total of 7 points)
c) Flexibility of parameters - the following parameters were considered: adjustable current intensity, adjustable session duration, adjustable number of sessions, adjustable electrode positioning (0 if no, 1 if minor adjustments, 2 if major adjustments were possible) - total of 5 points.
d) Ecological footprint - we evaluated the following aspects: batteries being rechargeable, batteries being replaceable, sponges being recyclable, sponges being reusable, devices being recyclable (total of 5 points).
e) Front-end interface - we considered the following grades, from 0 to 3: no interface, interface in the device itself, interface in the device itself, but with touch-screen, interface via smartphone app.
f) Data security - we considered the following aspects: compliance to GDPR, HIPAA and similar laws, mentioning of data encryption procedures, mentioning of data anonymization procedures, option for not collecting sensitive data (total of 4 points).

**Appendix 6. Steering Committee and Expert Panel Working Group**

**Supplementary Table 4. Authors included in the Steering Committee (SC) and the Expert Panel groups.** For the SC, 34 authors were invited and all agreed to participate. For the EP, out of 43 authors who were identified, we could not find the email address of one of them, 13 did not reply to our contact, and 2 declined to participate due to retirement or lack of time. Finally, 27 participants constituted the EP.

| Steering Committee (SC) (n=34) | Expert Panel (EP) (n=27) |
|---|---|
| Klaus Schellhorn | Isabela Bensenor |
| Marom Bikson | Shapour Jaberzadeh |
| Adam Woods | Donel Martin |
| Roy Hamilton | Leandro Valiengo |
| Abhishek Datta | Agnes Floel |
| Anna Wexler | Stephan Goerigk |
| Leigh Charvet | Christian Plewnia |
| Aurore Thibaut | Paul Fitzgerald |
| Walter Paulus | Leon Morales-Quezada |
| Felipe Fregni | Katia Monte-Silva |
| Marcel Simis | Paradee Auvichayapat |
| Erik Rehn | Filippo Brighina |
| Andrea Antal | Ulrich Palm |
| Wolnei Caumo | Dawson Kidgell |
| Ganesan Venkatasubramanian | Roi Kadosh |
| Lais Boralli Razza | Steven Laureys |
| Sarah H. Lisanby | Patrick Ciechanski |
| Colleen Loo | Mark George |
| Tarek Rajji | Vincent Clark |
| Chris Baeken | Kate Hoy |
| Zhi-De Deng | Michal Lavidor |
| Hamed Ekhtiari | Carlo Miniussi |
| Andre Brunoni | Marine Mondino |
| Jerome Brunelin | Alexander Okano |
| Flavio Frohlich | Claudia Oliveria |
| Ester Miyuki Nakamura Palacios | Rudi Raedt |
| Michael Nitsche | Dirk Ridder |
| Frank Padberg | ---- |
| Marie-Anne Vanderhasselt | ---- |
| Ines Violante | ---- |
| Paulo Boggio | ---- |
| Sandra Carvalho | ---- |
| Jorge Leite | ---- |

| | |
|---|---|
| Sven Vanneste | ---- |

**Appendix 7. Demographic characteristics of the included authors**

The ages of the panelists ranged from 25 to 69 years, most of them aged between 40 to 49 years (44% of the SC, and 33% of the EP). Participants resided in several countries, mostly in the US (SC n=11, EP n=3), followed by Brazil (SC n=6, EP n=5) and Germany (SC n=5, EP n=4). Regarding gender, 30% were female (SC n=10, EP n=8). Ninety-five percent of the participants hold a MD, PhD, or MD/PhD degree (SC n=31, EP n=27), and 78% of them had more than 10 years of experience in the field of non-invasive brain stimulation (SC n=26, EP n=22). Only 15% of members in the SC and 18% of members in the EP report did not have an ongoing tES trial with digital features, while the others reported having 1 or more ongoing tES trials with digital features. In total, 51 out of 61 participants reported that they are currently the PI of at least one ongoing tES trial with digital features.

**Supplementary Table5. Demographic and academic information for the steering committee (SC) and expert panel (PC)**

| | *Steering Committee (n=34)* | *Expert Panel (n=27)* |
|---|---|---|
| **Gender** | | |
| Male | 23 | 18 |
| Female | 10 | 8 |
| Other | 1 | 1 |
| **Age (years)** | | |
| 25-30 | 2 | 1 |
| 31-39 | 7 | 3 |
| 40-49 | 15 | 9 |
| 50-59 | 7 | 8 |
| 60-69 | 2 | 6 |
| Prefer not to say | 1 | 0 |
| **Highest Academic Degree** | | |
| Bachelor of Science | 1 | 0 |
| Master of Science | 2 | 0 |
| Doctor of Medicine (MD) | 6 | 6 |
| Doctor of Philosophy (PhD) | 19 | 16 |
| Doctor of Medicine and Philosophy (MD, PhD) | 6 | 5 |
| **Country of Residence** | | |
| Australia | 1 | 5 |
| Belgium | 3 | 1 |
| Brazil | 6 | 5 |
| Canada | 1 | 2 |
| France | 1 | 1 |
| Germany | 5 | 4 |

| | | |
|---|---|---|
| India | 1 | 0 |
| Ireland | 1 | 0 |
| Israel | 0 | 1 |
| Italy | 0 | 2 |
| New Zealand | 0 | 1 |
| Portugal | 2 | 0 |
| Sweden | 1 | 0 |
| Thailand | 0 | 1 |
| United Kingdom | 1 | 1 |
| United States | 11 | 3 |
| **Primary Field of Research (Except Non-Invasive Brain Stimulation)** | | |
| Biomedical Engineering | 2 | 0 |
| Cognitive Science | 2 | 2 |
| Computational Modeling | 1 | 0 |
| Ethics and Health Policy | 1 | 0 |
| Epidemiology | 0 | 1 |
| Neurology | 4 | 3 |
| Neuroscience | 3 | 6 |
| Neurosurgery | 0 | 1 |
| Pharmacology | 1 | 0 |
| Psychiatry | 14 | 11 |
| Psychology | 6 | 3 |
| **Primary Place of Work** | | |
| Private Company | 3 | 0 |
| Hospital | 12 | 8 |
| Research Institute (Government) | 2 | 0 |
| Research Institute (Independent) | 1 | 0 |
| University | 16 | 19 |
| **Experience in Non-Invasive Brain Stimulation Field (years)** | | |
| ≤5 | 1 | 1 |
| 6–10 | 7 | 4 |
| >10 | 26 | 22 |
| **Experience in conducting tDCS or rTMS clinical trials (years)** | | |
| ≤5 | 6 | 2 |
| 6–10 | 10 | 8 |
| >10 | 18 | 16 |
| No Response | 0 | 1 |
| **Experience in conducting tDCS or rTMS clinical trials using digital features (years)** | | |

| | | |
|---|---|---|
| ≤5 | 22 | 23 |
| 6–10 | 5 | 0 |
| >10 | 5 | 4 |
| No Response | 2 | 0 |

**Supplementary Figure 2. Geographical distribution of Delphi panel members (n=61; Steering committee=34 and Expert panel = 27).**

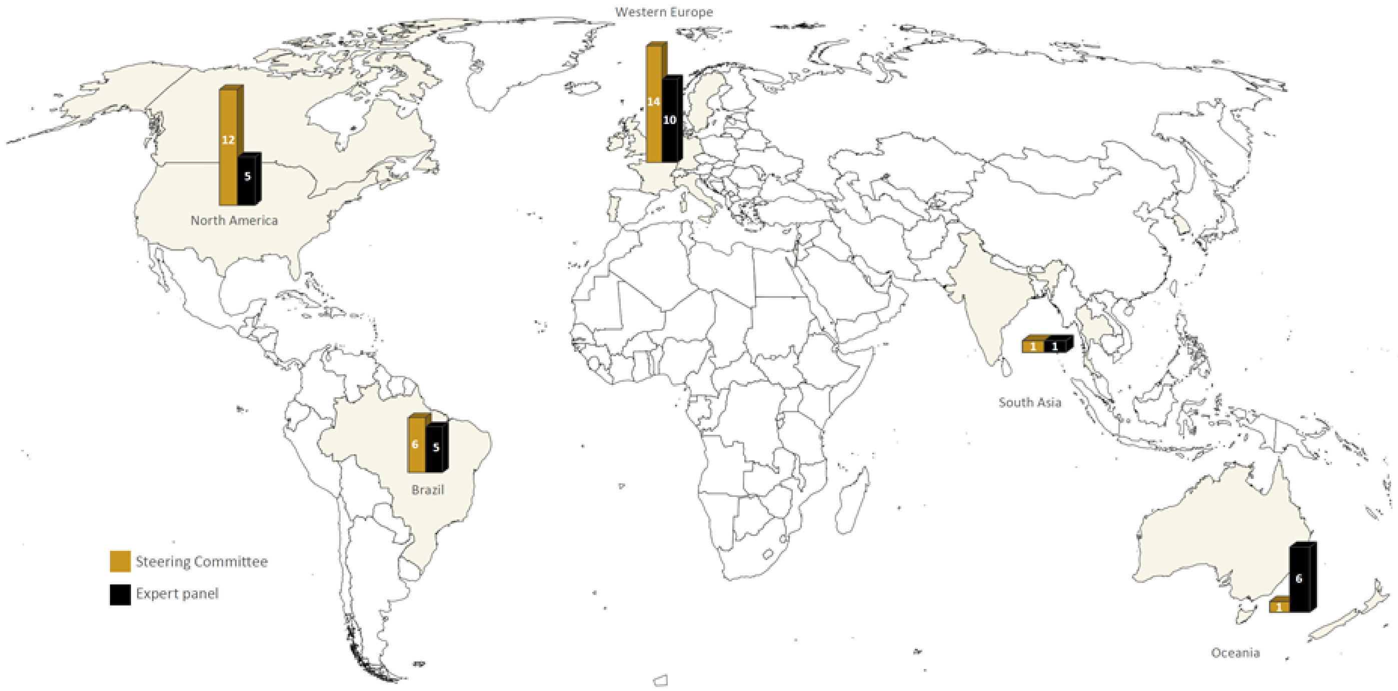

**Appendix 8. Systematic scoping review results**

Our search yielded 443 articles, of which 409 were excluded for several reasons, such as not tES studies, animal model studies, original articles, study protocol and other reviews (meta-analysis, systematic reviews, umbrella reviews and literature reviews) (**Supplementary Figure 3**). Therefore, 34 articles met the inclusion criteria and were selected for our scoping review. Of those, there were 9 recommendation articles [5–13], 10 guidelines [14–23], 10 critical reviews [24–33], and 5 expert consensus articles [12,23,34–3637–41]. This literature was used as the basis for discussion of the results obtained from the Delphi Panel.

**Supplementary Figure 3. Flowchart of the scoping review selection process**

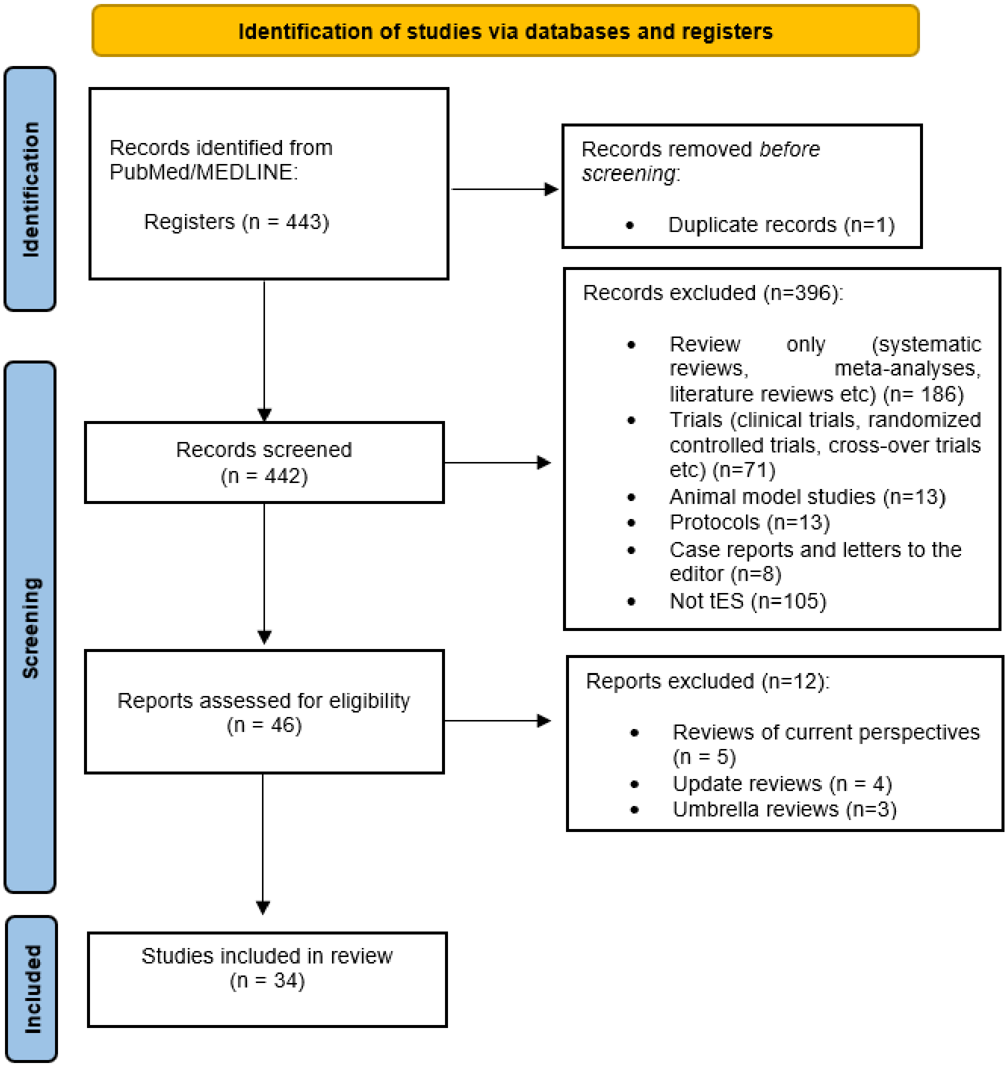

**Appendix 9. Rating scores for the SWOT assessment.**

**Supplementary Table 6. Ratings for "strengths" and "opportunities" of ptES clinical trials.** Ratings for items (scores 1-5, 1: strongly disagree, 2: disagree, 3: undecided, 4: agree, 5: strongly agree) are reported as mean (standard deviation) as well as the degree of agreement for each item as percentage. This table represents the ratings of 55 participants (24 from the steering committee and 31 from the expert panel).

| Questions | Agreement Level | | | | | |
|---|---|---|---|---|---|---|
| | Mean (SD) | Strongly Disagree | Disagree | Undecided | Agree | Strongly Agree |
| 1.1. Safety in Remote Use | 3.82 (1.14) | 7.3% | 9.1% | 3.6% | 54.6% | 25.5% |
| 1.2. Simple to Self-Perform tDCS | 3.78 (1.06) | 3.7% | 11.1% | 13.0% | 48.2% | 24.1% |
| 1.3. Being a Non-Pharmacological Intervention | 4.36 (0.8) | 1.8% | 1.8% | 3.6% | 43.6% | 49.1% |
| 1.4. Being a Non-Expensive Intervention | 4.13 (0.86) | 0.0% | 9.1% | 3.6% | 52.7% | 34.6% |
| 2.1. Being Scalable/ Having Low Marginal Costs | 3.96 (1) | 0.0% | 14.6% | 7.3% | 45.5% | 32.7% |
| 2.2. Association with Telemedicine | 4.45 (0.6) | 0.0% | 0.0% | 5.5% | 43.6% | 50.9% |
| 2.3. Possibility of Collecting Biobehavioral Data | 4.36 (0.65) | 0.0% | 1.8% | 3.6% | 50.9% | 43.6% |
| 2.4. Possibility of Combining Interventions | 4.51 (0.66) | 0.0% | 0.0% | 9.1% | 30.9% | 60.0% |
| 2.5. Possibility of Automate Procedures | 4.06 (0.9) | 0.0% | 5.6% | 20.4% | 37.0% | 37.0% |
| 2.6. Using 5G / IoT to Enhance ptES Use | 4.05 (0.78) | 0.0% | 3.6% | 16.4% | 50.9% | 29.1% |
| 2.7. Use of Design Thinking Strategies | 4.09 (0.75) | 1.8% | 0.0% | 12.7% | 58.2% | 27.3% |

Abbreviation: IoT, Internet of Things; ptES, portable transcranial electric stimulation; tDCS, transcranial direct current stimulation; SD, standard deviation.

The main identified **strengths** for using tES in the context of digital trials were:

(a) **Safety.** As digital trials are carried out mostly outside hospital settings, safety is a fundamental issue. Several reviews have shown that tES is safe, well-tolerable, and devoid of serious adverse events even with multiple repeated sessions of stimulation, when standard protocols are used [23,36,42,43 44]. Although it is associated with common adverse events, such as tingling, itching, and skin redness [45], these are short-lived, mild, and well-tolerated [23]. Nonetheless, not all tES devices might be equally safe for remote use, and some panelists expressed concern on the unsupervised use, using tES beyond recommendation guidelines, and unknown risks and adverse effects when applying tES in neuropsychiatric samples that have not yet been sufficiently studied. Another issue is that adverse events might vary between different tES devices, especially considering whether they were approved as certified, medical-grade materials or consumer devices that may not have these assurances.

**(b) Simplicity**. The skills necessary for electrode montage and session activation vary according to the tES device. However, since they are usually developed for the end-user, they should focus on simplicity, with clear instructions on how to perform each phase of the process. However, concerns were raised regarding the setup being used (e.g., some montages such as prefrontal bilateral stimulation would be simpler to use compared to motor cortex / extracephalic electrode placement).

**(c) Non-pharmacological intervention.** As pharmacotherapies frequently have adverse events that limit their use [46], present several contraindications, particularly in those with clinical comorbidities [47] and face also potentially hazardous drug-drug interactions, tES as a non-pharmacological intervention was seen as an appealing strength.

**(d) Affordable.** Traditional tES devices are relatively simple to be built in terms of electric engineering [48]. However, portable devices can be more technology-intensive, as a user-friendly interface contains microprocessors, bluetooth and wireless technologies, disposable sponges, rechargeable, customized batteries, programming, and continuous updates. Nonetheless, recent technological developments allowed companies to build tES devices with most of these features at relatively reduced costs. Still, costs vary according to the device and company, and they might be prohibitive in low-/middle-income countries or if insurance coverage is not provided (the current scenario in most countries). It was also mentioned that personnel costs (e.g., remote technicians who provide supervision or troubleshooting) should be factored into total costs.

Finally, the presence of positive pilot and pivotal RCTs was initially presented as a strength due to findings stating probable evidence of efficacy for several neuropsychiatric disorders [16,18,49]. However, most specialists pointed out that negative results have also been reported and the number of pivotal trials is still low. Here, other reported issues were that there are neither post-marketing approval tES studies nor controlled trials using tES. Finally, other strengths that did not reach consensus level were the possibility of using tES for cognitive enhancement; the design of tES devices being suitable for clinical research; and a potential large placebo effect, which could boost clinical outcomes or, conversely, lower effect size of the active-to-sham difference.

The main identified **opportunities** for using tES in the context of digital trials were:

**(a) Scalability.** In on-site tES trials, the main factors that keep recruitment and participation rates low are the space and personnel necessary to deliver tES sessions, and commute time of participants. Such constraints do not exist in tES digital trials, as sessions are self-delivered outside the research center. Therefore, a research assistant using the proper digital resources can simultaneously monitor and assist dozens of participants. In this context, digital trials are also scalable in space: while an on-site trial is restricted to a research centers' catchment areas, the catchment area of digital trials is undefined. In fact, spatial restrictions of digital trials are not *per se* physical, but mainly involving regulatory issues that might vary in different states or countries. Notwithstanding, if such issues are handled, a single center can coordinate a digital trial with an enrollment area as large as an entire country, sparing the need of multiple centers and therefore greatly reducing costs - an aspect that could be also leveraged internationally. However, lack of clear clinical indications might limit the likelihood of realizing the potential of scalability for the technology relative to clinical applications.

**(b) Telemedicine / Telehealth.** During the COVID-19 pandemic, teleconsultations have been widely adopted. For instance, in October 2020, 41% of psychiatric consultations in the US were done via telemedicine vs. < 1% before the pandemic [50]. Although it is still unclear whether telemedicine will still be reimbursed after the pandemic, the "telemedicine mindset" of patients and physicians seems to have changed from an exception to another alternative in medical care, that could be even preferable in some contexts (e.g., rural patients, short appointments) [50]. This "mindset shift" is beneficial for digital trials in which most or all clinical assessments are performed online. Likewise, as pharmacy delivery services are being discussed, tele-psychotherapy being more adopted, and digital mental health apps use increased during the pandemic, tES as a home-use treatment modality might be increasingly accepted [51].

**(c)** Employment of mobile Health technologies**,** particularly considering (1) digital phenotyping since, as a wearable digital assessment can also be performed by using apps and tES can leverage digital assessment collection using scalp biosensors integrated to the electrodes, and (2) combination with digital interventions, such as apps designed for online cognitive training, mindfulness/meditation, or psychotherapy, possibly enhancing overall efficacy [52] and also standardizing tES administrations by reducing the influence of the environment during the session.

**(d) Automatization.** Processes of digital trials can be partially automated at several steps (see SIPOC below). For instance, (1) volunteers can fill out electronic forms and be automatically excluded according to eligibility criteria; (2) potential participants can be scheduled and contacted automatically (e.g., RedCap [53], a popular academic data platform, has scheduling and text messaging / e-mail sending modules) for online screening (a step that can be enriched by using machine learning algorithms to predict the likelihood ratio to be recruited); (3) enrolled participants can provide digital consent, have tES devices shipped to their homes, and being instructed how to use it via videos and/or augmented reality techniques; and (4) interaction with the staff can be mediated by psychiatric chatbots [54] for filtering between simple questions, reporting of adverse events and need of medical care. However, home use would still depend on someone handling the system and professionals doing supervision. In fact, patients might get enhanced benefits with periodic live connections with the staff, even if briefly and remotely.

**(e) 5G / Internet of Things.** 5G is an emerging technology of mobile communication that is being implemented worldwide. Due to its increased bandwidth, very high data transmission, ultra low latency, and massive network capacity, a fully-implemented 5G network could allow seamless device-to-device communication ("Internet of Things") [55]. This will offer enormous possibilities for connecting tES with other devices that can instantly assess digital phenotypes [56] and provide readouts. Also due to 5G connectivity, such multidimensional granularity could be processed by cloud-based systems in a matter of milliseconds, eventually allowing the development of closed-loop systems [57] for tES. Notwithstanding, not everyone would benefit from these advancements, such as people living in areas with no 5G technology. In addition, the present generation of tES devices does not have such technologies.

**(f) Design thinking**. Although increasingly simple to operate, some aspects of tES devices could still be improved to be centered around the patients´ perspectives [58]. For instance, tES devices could be adapted for those who are physically or cognitively impaired to guarantee ease of use in such cases, or modified to look less "medical" and be used in more ecological, real life settings. However, this opportunity requires collaborations between tES companies, academics, and other stakeholders such as patients using these devices and providing feedback.

**Supplementary Table 7. Ratings for "threats" and "weaknesses" of ptES clinical trials.** Ratings for items (scores 1-5, 1: strongly disagree, 2: disagree, 3: undecided, 4: agree, 5: strongly agree) are reported as mean (standard deviation) as well as the degree of agreement for each item as percentage.

| Questions | Agreement Level | | | | | |
|---|---|---|---|---|---|---|
| | Mean (SD) | Strongly Disagree | Disagree | Undecided | Agree | Strongly Agree |
| 1.1. Potential Cyber-Violations (Hacking) | 3.49 (0.9) | 1.8% | 12.7% | 29.1% | 47.3% | 9.1% |
| 1.2. Difficulties in Keeping Sensitive Data Safe (Confidentiality) | 3.35 (1.02) | 3.6% | 21.8% | 18.2% | 49.1% | 7.3% |
| 1.3. Use by Lay People (DIY / Recreational use) | 3.98 (1.05) | 1.8% | 9.1% | 16.4% | 34.6% | 38.2% |
| 1.4. Unclear Regulatory Status | 3.73 (0.95) | 1.8% | 10.9% | 18.2% | 50.9% | 18.2% |
| 2.1. Difficulties with Remote Supervision | 3.84 (0.92) | 1.8% | 9.1% | 12.7% | 56.4% | 20.0% |
| 2.2. Bioethical Issues | 3.45 (1.03) | 1.8% | 20.0% | 23.6% | 40.0% | 14.6% |
| 2.3. Difficulties with Proper Electrode Positioning | 3.72 (0.92) | 0.0% | 13.0% | 20.4% | 48.2% | 18.5% |
| 2.4. Skin Burns as a Relevant Side Effect | 3.2 (0.95) | 1.8% | 25.5% | 29.1% | 38.2% | 5.5% |
| 2.5. Low Clinical Evidence of Efficacy | 3.18 (1.17) | 10.9% | 18.2% | 21.8% | 40.0% | 9.1% |
| 2.6. Higher Rates of Serious Adverse Events | 3.11 (1.12) | 3.6% | 32.7% | 25.5% | 25.5% | 12.7% |

Abbreviation: DIY, do it yourself; ptES, portable transcranial electric stimulation; SD, standard deviation.

The main identified **weaknesses** for using tES in the context of digital trials were:

**(a) Insufficient or absent supervision.** Concerns were identified by the panelists regarding patients themselves manipulating the device and lack of and difficulties in adequate, proper supervision and monitoring, compared to on-site clinical trials. This could lead to misuse (i.e., over- or under-dosing, particularly if users need to set the parameters themselves); diversion of the device (i.e., device being used by someone other than the patient with risks of injuries or serious adverse events); or device being used outside of the medical context (i.e., for gaming, performance enhancement, or military applications). The panelists also commented that there could be issues in the reproducibility of findings as, without monitoring / supervision, there is no assurance that the use was as instructed and intended.

**(b) Difficulties in obtaining accurate adequate placement** - tES positioning systems are derived from anatomical scalp measurements that might be suboptimal, as electric field modeling simulations showed that there is large variability of cerebral electric field distribution across patients using the same tES parameters, and that such variation can influence outcomes [59]. In addition, even small changes of electrode orientation (horizontal or vertical) or small drifts from the target location can potentially impact outcomes [40 60]. However, experts considered that this is a greater risk for non-experienced users, which could be reduced via training, guidelines, standardization, and continuous improvement of tES devices. In this context, companies should develop and test new methods for assuring correct placement of electrodes (e.g., [61]).

Several other potential weaknesses did not reach consensus during the process of rating and validation. For instance, some panelists raised concerns related to bioethics**.** The principle of justice could be violated in tES, as treatment would be offered only for those that have access to technology (internet connectivity, smartphones), some degree of digital literacy, and no physical/sensory impairments. Therefore, large groups such as those with lower socioeconomic status, populations from low-/middle- income countries, and older groups (in fact, groups with increased rates of neuropsychiatric disorders) would have restricted access to tES technology. Also, most of the panelists did not reach a consensus on weaknesses related to adverse events, such as skin burns, which could be an issue given that supervision levels are lower than in on-site trials. However, many experts did not consider this as an issue, as skin burns are not a common adverse event, which can be mitigated by training and standardization, and have not been reported to occur at-home [62]. Likewise, serious adverse events, such as manic switch and suicidal ideation, were not considered a weakness [24]. Finally, panelists were divided regarding whether low clinical efficacy is a weakness, as almost half of them indicated that there is lack of definite clinical evidence for most conditions [18], and high variability of effects in conditions with a good level of clinical evidence of efficacy [63].

Finally, the main **threats** for tES digital trials implementation were:

**(a) Do-it-yourself (DIY) and recreational use.** tES application without any professional guidance has been drawing attention and concerns from researchers and clinicians in the field for a while now, once the DIY-tES community has gained notoriety in the past years. The main concern raised is the fact that DIY-tES practice is particularly related to safety issues as it takes place outside of a clinical setting, commonly by naive users [7]. In this sense, the concern in harnessing the opportunity of tES use is the widespread consumer availability in the absence of careful dosing guidance outside of clinical care, since several tES companies have already launched 'direct-to-consumer' tES devices. Protocols simply using tES without paying attention to every detail of e.g., electrode montage may just induce opposite effects [64].

(b) **Regulatory status**. Medical devices have to undergo different regulation processes according to the state, country or region considered. In Europe, the new regulatory standards for medical devices [65] have increased the requirements for approval (CE certification). Non-medical brain stimulation devices (devices without an intended medical purpose) will have to meet similar requirements for safety and quality as medical devices. Previously, it was easier to obtain marketing approval in the EU by claiming equivalence with devices used in clinical trials, and own clinical data obtained by the manufacturer from randomized controlled phase II or III trials played almost no role. In the US, premarket reviews of low-/medium-risk medical devices are necessary before FDA approval, while device approval by similarity (the 510(k) clearance) of approved devices are common [66]. tES devices are not formally approved by the FDA, bringing a regulatory gap to their use: on one hand, some companies can commercialize devices under a "regulatory device exemption"; whereas iontophoresis devices that can be adapted to perform electrical stimulation (without the same safety standards of tES devices) are freely marketed [67]. In fact, more than 1000 different electrical devices have been cleared by FDA since 1977, for several reasons, one of the most common being "general wellness devices" [68]. Interestingly, most of these devices use parameters (e.g., dose and electrode size) that exceed safety guidelines of tES devices [68]. Portable tES devices, particularly those with systems that collect behavioral and biological data and perform wireless communications, pose additional challenges, as such devices can also be considered as mobile Health (mHealth) systems demanding specific regulations [69]. However, some specialists considered that the current regulatory status is not an inherent weakness of tES devices; instead, this is mainly an issue for device companies and those using tES beyond clinical research.

Several threats initially presented by the main authors and/or the SC did not reach further consensus. At the review phase by the SC, disruptions in the supply chain and ecological footprint were not considered threats by most members. Regarding the former, the main authors considered that large trials could need hundreds or thousands of devices that would need to be manufactured, shipped, and stored in advance - therefore, disruptions of the supply chain (as occurred during the pandemic) would delay respective studies. However, the SC considered that these issues were restricted to the early phases of the pandemic, and/or handled by adopting lean management models focused on pull production and waste reduction (for instance, the "Toyota model"). Regarding the latter, the main authors considered that the use of non-recyclable devices and disposable sponges could impact ecological sustainability, an increasing concern worldwide. However, most specialists stated that these issues are offset by the gains in avoiding daily transportation to the clinical center (compared to TMS or on-site tES) or in not manufacturing pills. Also, companies are increasingly abiding to the environmental, social, and governance criteria. During the grading phase, two potential threats were identified by most of the panel, but did not reach the 60% threshold. One of them was risks related to hacking and cyber-security**.** A recent systematic review showed that most mHealth apps are developed with insufficient cyber-security [70]. On the other hand, most companies stated that state-of-the-art encryption methods are being adopted. The other one was risks related to confidentiality and anonymity**,** considering risks related to collecting sensitive data, different data protection laws and collection of non-authorized data. On the other hand, several new data law protections (e.g., GDPR, LGPD, CCPA, and POPI) have been enforced to protect the use of sensitive and personal data by companies.

**Appendix 10. Rating scores for the SIPOC assessments**

**Supplementary Table 8. Ratings for "methodological perspectives" of ptES clinical trials.** Ratings for items (scores 1-5, 1: strongly disagree, 2: disagree, 3: undecided, 4: agree, 5: strongly agree) are reported as mean (standard deviation) as well as the degree of agreement for each item as percentage.

| Questions | Agreement Level | | | | | |
|---|---|---|---|---|---|---|
| | Mean (SD) | Strongly Disagree | Disagree | Undecided | Agree | Strongly Agree |
| 1.1. Can Use Different Study Designs | 3.91 (0.97) | 3.6% | 7.3% | 7.3% | 58.2% | 23.6% |
| 1.2. Can Perform Adaptive Trials | 3.85 (0.99) | 3.6% | 7.3% | 12.7% | 52.7% | 23.6% |
| 1.3. ptES Are Already Well Developed | 3.62 (0.93) | 1.8% | 12.7% | 20.0% | 52.7% | 12.7% |
| 1.4. Longer Follow-up | 4 (0.88) | 1.8% | 5.5% | 10.9% | 54.6% | 27.3% |
| 1.5. Can Recruit more Participants | 4.22 (0.76) | 0.0% | 5.5% | 3.6% | 54.6% | 36.4% |
| 1.6. Faster than On-Site | 3.75 (0.84) | 0.0% | 9.1% | 23.6% | 50.9% | 16.4% |
| 1.7. Greater External Validity | 3.6 (0.81) | 0.0% | 10.9% | 27.3% | 52.7% | 9.1% |
| 1.8. Greater Internal Validity | 2.8 (0.87) | 3.6% | 34.6% | 43.6% | 14.6% | 3.6% |
| 1.9. Can Collect more Data | 4.04 (0.86) | 0.0% | 9.1% | 7.3% | 54.6% | 29.1% |
| 1.10. Can Be Combined with Other Therapies | 3.6 (0.99) | 1.8% | 14.6% | 21.8% | 45.5% | 16.4% |
| 1.11. Is Scalable | 4.09 (0.8) | 0.0% | 3.6% | 16.4% | 47.3% | 32.7% |
| 2.1. Should Be Done Only after On-Site Studies | 3.44 (1.24) | 7.3% | 21.8% | 10.9% | 40.0% | 20.0% |
| 2.2. Need to Develop New Assessment Methods | 3.65 (0.87) | 0.0% | 10.9% | 27.3% | 47.3% | 14.6% |
| 2.3. Increased Risk for Suicidality | 3.2 (1) | 3.7% | 22.2% | 31.5% | 35.2% | 7.4% |
| 2.4. Increased Risk for Manic Switch | 3.27 (0.93) | 1.8% | 20.0% | 34.6% | 36.4% | 7.3% |
| 2.5. Increased Risk for Cognitive Effects | 3.09 (0.93) | 3.6% | 23.6% | 36.4% | 32.7% | 3.6% |
| 2.6. Increased Risk for SAEs | 2.94 (0.97) | 5.7% | 30.2% | 30.2% | 32.1% | 1.9% |
| 2.7. ptES Will Substitute On-Site | 2.53 (1.09) | 14.6% | 43.6% | 21.8% | 14.6% | 5.5% |
| 2.8. More Dropouts | 3.04 (0.98) | 3.6% | 29.1% | 32.7% | 29.1% | 5.5% |
| 2.10. More Randomization Biases | 2.89 (0.96) | 3.6% | 38.2% | 25.5% | 30.9% | 1.8% |
| 2.11. More Blinding Biases | 3.33 (0.92) | 3.6% | 18.2% | 21.8% | 54.6% | 1.8% |
| 2.12. More Sham Biases | 3.19 (1.08) | 5.6% | 27.8% | 14.8% | 46.3% | 5.6% |

Abbreviation: SAEs, serious adverse events; ptES, portable transcranial electric stimulation; SD, standard deviation.

**Supplementary Table 9. Ratings for "research team features" in ptES clinical trials.** Ratings for items (scores 1-5) are reported as mean (standard deviation) as well as the degree of agreement for each item as percentage

| Questions | Agreement Level | | | | | |
|---|---|---|---|---|---|---|
| | **Mean (SD)** | **Strongly Disagree** | **Disagree** | **Undecided** | **Agree** | **Strongly Agree** |
| 1. Expert in Digital Marketing | 3.61 (0.86) | 1.85% | 7% | 30% | 50% | 11% |
| 2. Expert in Data Science | 3.96 (0.79) | 0.00% | 4% | 22% | 49% | 25% |
| 3. Expert in Front-end Interface | 4 (0.77) | 0.00% | 4% | 18% | 53% | 25% |
| 4. Expert in Back-end Interface | 3.98 (0.89) | 0.00% | 7% | 18% | 44% | 31% |
| 5. Expert in Data Security | 4.25 (0.64) | 0.00% | 0% | 11% | 53% | 36% |
| 6. 24/7 Team Availability | 4.13 (0.94) | 0.00% | 9% | 11% | 38% | 42% |

Abbreviation: ptES, portable transcranial electric stimulation; SD, standard deviation.

We identified 4 main processes (recruitment, pre-screening, screening, and participation) to compare the processes in tES trials adopting or not digitalization. The panelists noted that trials will not likely be purely digital or analog, and different degrees of digital features can occur at each process. For instance, participants can be recruited through social media, but screened onsite. Another aspect is that digital processes would allow to collect and process more metrics, which can aid in adjusting processes (e.g., when using social media, an ideal number of volunteers per week can be achieved by using key performance indicators such as engagement, leads, clicks, conversions, impressions, and others). In addition, digital processes are overall quicker than non-digital ones - therefore, a larger number of participants could be recruited faster, and possibly followed-up for longer periods of time. Finally, another defining difference of digitalization processes is their potential of scalability due to the use of digital assessments and telemedicine. However, some experts noticed that, since most processes are relatively new, lack of rigor and reproducibility is a concern. Another potential issue is that attrition might be higher in digital trials, as the selected sample might have lower compliance, as the requirements for participating in the trial are lower, and be less motivated to participate, as engagement with the digital trial is more restricted. These aspects are discussed below.

**Methodological aspects**

We further divided this section into characteristics that did and did not reach consensus by the evaluation of the SC and EP members, along with qualitative aspects and comments.

**Consensus reached**

Regarding **study design,** tES digital trials are compatible with all study designs traditionally employed in on-site tES RCTs, such as parallel [71], factorial [72], non-inferiority [73], and cross-over [74] designs, as well as pilot and pivotal trials. Additionally, pragmatic trials - i.e., trials that test if the intervention works in the "real world", favoring external validity (i.e., few exclusion criteria, community interventions, tele-medicine, and longer follow-up periods) [75] - could be particularly employed in digital trials that are scalable and therefore with lower costs per patient [76]. Also, due to tES and digital scalability, multi-arm trials are an appealing design since several interventions can be tested at once. However, within-subject designs - i.e., studies in which patients are assigned to several intervention groups over time - might be more complex as this would require greater remote experimental manipulation. Moreover, for this particular design and depending on the details of active and sham stimulation, the participants could guess their assigned group as the rate of adverse events might be different according to current intensity. Some complex adaptive clinical trial designs can also be adopted in tES digital trials. The adaptations might include sample size re-estimation, stopping for futility, changing allocation ratio between groups, dropping or adding new groups, modifying treatment arms or seamlessly "upgrading" to the next clinical phase [77] - considering that tES devices present technologies that allow for most of these adjustments to be performed remotely. As adaptive designs require updating information as the trial progresses, digital trials are advantageous since computing and adaptation can be performed automatically. However, not all adaptive designs are straightforward to be performed in tES trials. For instance, modifying treatment arms might not be possible, especially if this involves electrode replacement or dosage adjustment that would require on-demand hardware and/or software adjustments - although technological progress will make this possible in the future.

Due to the need of daily visits to the research center, on-site tES trials face difficulties in recruiting large samples [35]. Moreover, they present some degree of selection bias, as enrolled participants are those who have the availability for performing daily visits to the research center, such as those with higher income, living near the research center and with characteristics that might be different than the target sample (e.g., patients with negative symptoms of schizophrenia, depression, or stroke might have lower severity than those patients who cannot commute to the clinical center). Therefore, tES trials present lower recruitment restrictions compared to on-site trials performed in clinical centers and, since the sessions are self-applied, this allows for the recruitment of larger and more diverse samples, increasing the pool of available participants, and theoretically increasing the external validity due to scalability. Also, scalability would allow recruitment of more participants and perform trials faster than on-site tES trials.

On the other hand, many panelists noted that external validity can be also an issue in tES digital trials, as the selected sample could present higher digital literacy, lower age (for instance, older groups are more likely to have some degree of physical or sensory impairment, making the handling of tES devices more difficult) and more participants that live in areas with internet connectivity. To evaluate tES effects in these groups, it would be necessary to adapt the device, provide smartphones, tablets, pads, and/or perform community-based clinical trials. Also, if recruitment strategies are performed solely using social media, the trial results would have restricted generalizability for people that do not use such media, and the trials might only select those prone to use the intervention. This issue would be partially overcome if recruitment is based on socio-demographic characteristics, with the aid of similar technologies used for targeted digital marketing campaigns. Nonetheless, the external validity of traditional tDCS trials is also substantially affected by the need to come to the stimulation center for several sessions.

Combining tES with cognitive, motor, or psychological interventions can boost neuroplasticity and enhance treatment efficacy (for a review, see [52]). Considering that tES is self-delivered, participants can use it in a diverse range of settings - for instance, coupled with physical therapies or digital apps that deliver cognitive-based or psychologically-based applications. Another advantage of combining tES with simultaneous behavioral interventions is that, if this is not done, participants will necessarily engage in other cognitive activities while at home - as a result, a plethora of "brain states" would be stimulated between and within-subjects over the course of a trial, which could result in larger variability of effects. Ideally, all participants should be engaged in a uniform simultaneous activity- even if that activity is not designed to be therapeutic itself.

Combining tES with concomitant interventions was perceived as an advantage that reached consensus by the panelists. However, they also noticed that there might be issues regarding: (1) checking adherence of the concomitant intervention, which would eventually require facial recognition or verifying user logins, which would be challenging if the intervention is promoted by a third-party; (2) controlling engagement with the digital intervention is also challenging and could be measured by using performance metrics and gamification features; (3) performing blinding and placebo of the concomitant intervention, if required by study design; and (4) most tES systems are not yet programmed for a straightforward experience of concomitant tES activation and digital intervention, requiring participant's pairing for each session (if an "online" intervention is desired). All these issues can increase variability of effects and decrease reliability of findings; therefore, although exciting, this combined approach might require further technological development and methodological validation.

Finally, panelists considered that, overall, tES devices are already well-developed and performing digital trials using this technology is feasible.

**No consensus reached**

The panelists were divided on whether greater internal validity can be achieved in tES digital trials. Internal validity is low when biases occur. We asked the panelists about several sources of bias and in many aspects the proportions of agreement and disagreement were around evenly distributed, with many comments provided by the panelists that are described in this section.

Although randomization procedures are essentially similar in on-site and digital tES trials (i.e., based on a randomized, computer-generated list), allocation procedures are different. Large on-site trials often use sealed, opaque envelopes containing the code corresponding to the group assigned for each participant [78–80]. Theoretically, this method could also be employed in digital tES trials as codes can be assigned to sham or active tES devices (i.e., "in the hardware") that would be shipped to patients. However, digital trials also bring the opportunity for seamless randomization-allocation procedures ("in the software"). For instance, Soterix Medical tES devices can be activated by a website- or SMS-delivered activation code that allocates participants to the randomized group. Flow devices are activated via a Bluetooth connection between the device and its app. In this case, allocation occurs as the patients login. However, both digital procedures can be vulnerable to contamination bias [81], i.e., the receipt of active treatment in those receiving control treatment. For instance, if the SMS-code does not have a short activation period, it could be inadvertently used by other participants (for instance, by posting the SMS-code on the Web) - a similar issue could occur if logins and passwords are shared online. A solution to this risk is allotment of activation codes to specific devices and/or with time limits. For home-based trials there is also the risk that the user may be substituted. To address this, activation could require biometry, facial recognition, 2-step confirmation, or periodic online verification by the study team. In theory, apps can be hacked and modified to deliver active and not sham stimulation or, even, hacking of the central server (or servers that have copies of the randomization list) might expose the entire list of participants of the study, reinforcing the need for companies to employ state-of-the-art security procedures.

Study blinding (or masking) is a set of procedures used to keep participants and researchers unaware of the allocation group. In tES trials, blinding effectiveness is essentially related to the parameter space of the control group relative to the test group. For instance, blinding in trials comparing two active doses of tES (e.g. 1 vs. 2 mA) might be easier to achieve than comparing different electrode montages that would be readily visible to the participants and to the staff. Notwithstanding, the most common comparison in tES trials is between an active and a sham (current ramp up/down) group [82]. Although the use of sham tES procedures has been debated [12,32]; the reliability of blinding by sham tES will depend strongly on the sensation in the active arm, which in turn depends on factors such as electrode design, ramp style, and operator training [83,84]. Other factors such as subtle investigator priming could have been involved in blinding breaking [85]. Nonetheless, most studies challenging blinding effectiveness used single-session, cross-over designs and/or volunteers that were non-naive to tES [12]; in contrast, large tES trials demonstrated that sham tES is as effective as a placebo-pill - the "gold standard" in RCTs - in maintaining blinding integrity [86]. Digital trials using tES might maintain or even improve blinding effectiveness, considering that participants will perform the sessions at home, with no contact between researchers and participants. Moreover, more flexibility in testing the parameter space would be allowed. Nonetheless, there are risks of blinding breaking as patients can manipulate the tES devices - for instance, by using amperemeters to measure the current between electrodes, by taking pictures of the montage and posting them on the Web, or by discussing and sharing their experiences in several social media venues (e.g., Reddit, Instagram, among others). We recommend that informed consent states that neither manipulation of the devices nor sharing pictures of them are allowed. It is also possible to model this statistically by integrating the patients' guesses regarding the intervention [87]. On the other hand, it is unclear how likely this would pragmatically occur, warranting further investigation. Some of these issues also affect traditional tES trials.

Sham stimulation is the placebo/control condition used in many neuromodulation studies. In tES trials, sham stimulation includes a brief active current applied to mimicking typical initial sensations such as itching and tingling caused by the active protocols in order to keep participants and personnel blinded to the administered intervention [84]. The optimal sham tES protocol is still being discussed in the literature [84] and tES devices usually allow researchers to tailor their own sham protocols. For tES digital trials, sham stimulation can be automated and delivered according to the code allocating the participant to the randomized group. We verified that the same methods (for instance, ramp-up/ramp-down periods, or a very low active current) employed in traditional tES can be used with tES devices. Information for participants should be kept to a minimum to avoid identification of sham.

Regarding assessments**,** digital trials permit greater flexibility in collecting data through using several questionnaires that can be applied remotely through the Web or by using apps. Questionnaires developed to be applied by the clinician can be used in teleconsultations, although construct validity when using them in digital contexts might not be similar and can require revalidation. More importantly, digital trials can leverage data collection by using mobile technologies that permit digital phenotyping using ecological momentary assessments, meta-data information, and physiological and activity data that can be captured by mobile phones [76,88], such as digital trackers for activity levels, sleep, facial expression, voice recording, natural language processing, and social sensing (i.e., quantifying social media activity). Also, gamified apps can be used to collect cognitive data. As tES is also a wearable device, technical developments could allow additional data collection such as EEG recording, heart rate and its variability, skin conductance, and other information that can be extracted through sensors placed over the skin. Also, digital assessments of **severe, serious or unexpected adverse events** (e.g., suicidality, manic switch, hazardous cognitive effects, and others) might be more complex remotely, and would require specific technological development - in fact, similarly to pharmacotherapy, as drug treatment also occurs remotely. On the other hand, granular data collection has several issues. For instance, there are greater challenges regarding data security, privacy, and anonymity. Thus, all collected data must be strictly necessary to the trials' aims, previously authorized by the participant, and stored in cyber secure servers that preferentially employ blockchain technologies [76].

Attrition rates might be lower or higher compared to digital tES trials than on-site trials that are around 6% [42]. On the one hand, attrition related to commuting to the clinical center would be null. On the other hand, participants would have less opportunities to discuss and interact with the research team, and this reduced contact could decrease motivation to participate in the study. Moreover, the selected sample in digital trials could be less committed than samples participating in on-site trials that decided in advance to invest time and effort in participating in the study. Therefore, tele-monitoring and proper interaction with participants, managing their expectations and credibility of the team, and reinforcing the need to abide by the study protocol, would be essential to decrease attrition. Finally, complex instances of non-adherence might be relevant: for instance, participants might give their devices to third parties to stimulate themselves, or they might connect the electrodes to saline solutions or wires that conduct electric current, mimicking a stimulation session. In such cases, using resources such as facial recognition would be necessary.

Finally, almost 60% of the panel (but still below the threshold) considered that tES should be performed only after on-site trials for a new condition. Likewise, between 50%-60% of the panel rejected the statement of tES trials substituting on-site trials. Taken together, these findings indicate that it is still unclear which role tES digital trials will serve in the near future. But it is also possible that results from digital trials with tES will change this scenario.

**Appendix 11. Rating scores for general recommendations**

**Supplementary Table 10. Ratings for general recommendations on digitizing ptES clinical trials.** Ratings for items (scores 1-5, 1: strongly disagree, 2: disagree, 3: undecided, 4: agree, 5: strongly agree) are reported as mean (standard deviation) as well as the degree of agreement for each item as percentage.

| Questions | Agreement Level | | | | | |
|---|---|---|---|---|---|---|
| | Mean (SD) | Strongly Disagree | Disagree | Undecided | Agree | Strongly Agree |
| 1. Develop Better Methods for Randomization and Allocation | 3.94 (0.81) | 0.0% | 3.7% | 24.1% | 46.3% | 25.9% |
| 2. Develop Better Methods for Sham Stimulation | 4.04 (0.7) | 0.0% | 1.9% | 16.7% | 57.4% | 24.1% |
| 3. Develop Methods to Decrease Attrition | 4.11 (0.63) | 0.0% | 1.9% | 9.3% | 64.8% | 24.1% |
| 4. Enhance External Validity | 4 (0.67) | 0.0% | 3.6% | 10.9% | 67.3% | 18.2% |
| 5. Combine ptES with Concomitant Interventions | 4.4 (0.56) | 0.0% | 0.0% | 3.6% | 52.7% | 43.6% |
| 6. Collect Ecological Data | 4.18 (0.58) | 0.0% | 0.0% | 9.1% | 63.6% | 27.3% |
| 7. Develop Safe Methods to Store Data | 3.71 (1.01) | 1.8% | 10.9% | 25.5% | 38.2% | 23.6% |
| 8. Develop Methods to Assess AEs | 3.6 (0.95) | 0.0% | 14.6% | 29.1% | 38.2% | 18.2% |
| 9. Develop Methods to Assess SAEs | 3.84 (0.81) | 0.0% | 5.5% | 25.5% | 49.1% | 20.0% |
| 10. Develop Methods to Assess Unexpected AEs | 3.84 (0.74) | 0.0% | 3.6% | 25.5% | 54.6% | 16.4% |

Abbreviation: SAEs, adverse events; ptES, portable transcranial electric stimulation; SD, standard deviation.

#### Recommendations

##### For the team performing digital tES trials

The panelists agreed that teams performing tES digital trials should have at least one person specialized in: (1) digital marketing strategies to aid in online recruitment; (2) data science and visualization; (3) design thinking for enhancing user experience via development of front-end features (although it was noticed by some experts that this matter mainly concerns companies); (4) app programming for enhancing user experience via development of back-end features; (5) a specialist on issues related to data security, integrity, anonymity, and replicability [8,9]; and (6) that the team should be always ("24/7") available. The feasibility of these recommendations should be further debated, as they would require more resources. In fact, most recommendations here fit with companies and for pivotal studies, and not necessarily for scientists running pilot studies that would not have all the capabilities mentioned above.

On the other hand, there was no majority to the statements that people skilled in machine learning techniques for developing predictive algorithms are necessary, as this depends on the trial goals. Also, most of the SC disagreed that "digital trials would require fewer clinicians, as part of the evaluations would be automated or collected without the need of raters".

##### For further research

Interestingly, despite a lack of consensus on whether tES digital trials present several types of biases, most of the panelists recommended that further research should be carried out regarding development of better methods for (a) randomization / allocation, and (b) sham methods. Also, further research was recommended to develop or refine methods to enhance (c) adherence and (d) external validity of the trials. It was also stated that more research should be devoted to aspects such as (e) combination of interventions, (f) biobehavioral data collection, (g) enhanced data security, and methods to assess (h) serious and (i) unexpected adverse events (but not adverse events in general). Finally one possibility is that hybrid trials with an online component can address some of the weaknesses identified here. For instance, the subject may be trained onsite and then do sessions online and/or assessments (or part of them) may be done online.


1 Teoli D, Sanvictores T, An J. SWOT Analysis. In: StatPearls. Treasure Island (FL): StatPearls Publishing, 2020.

2 Brown C. Why and how to employ the SIPOC model. *J Bus Contin Emer Plan* 2019; **12**: 198–210.

3 Akins RB, Tolson H, Cole BR. Stability of response characteristics of a Delphi panel: application of bootstrap data expansion. *BMC Med Res Methodol* 2005; **5**: 1–12.

4 Pham MT, Rajić A, Greig JD, Sargeant JM, Papadopoulos A, McEwen SA. A scoping review of scoping reviews: advancing the approach and enhancing the consistency. *Res Synth Methods* 2014; **5**: 371–85.

5 Zhang Y, Song H, Chen Y, Zuo L, Xia X, Zhang X. Thinking on Transcranial Direct Current Stimulation (tDCS) in Reading Interventions: Recommendations for Future Research Directions. *Front Hum Neurosci* 2019; **13**: 157.

6 Thibaut A, O'Brien AT, Fregni F. Strategies for replacing non-invasive brain stimulation sessions: recommendations for designing neurostimulation clinical trials. *Expert Rev Med Devices* 2017; **14**: 633–49.

7 Sierawska A, Prehn-Kristensen A, Moliadze V, *et al.* Unmet Needs in Children With Attention Deficit Hyperactivity Disorder—Can Transcranial Direct Current Stimulation Fill the Gap? Promises and Ethical Challenges. *Front Psychiatry* 2019; **10**: 334.

8 Kim D-J, Moussa-Tooks AB, Bolbecker AR, *et al.* Cerebellar-cortical dysconnectivity in resting-state associated with sensorimotor tasks in schizophrenia. *Hum Brain Mapp* 2020; **41**: 3119–32.

9 Fried PJ, Santarnecchi E, Antal A, *et al.* Training in the practice of noninvasive brain stimulation: Recommendations from an IFCN committee. *Clin Neurophysiol* 2021; **132**: 819–37.

10 Fregni F, Nitsche MA, Loo CK, *et al.* Regulatory Considerations for the Clinical and Research Use of Transcranial Direct Current Stimulation (tDCS): review and recommendations from an expert panel. *Clin Res Regul Aff* 2015; **32**: 22–35.

11 Deer TR, Mekhail N, Provenzano D, *et al.* The appropriate use of neurostimulation of the spinal cord and peripheral nervous system for the treatment of chronic pain and ischemic diseases: the Neuromodulation Appropriateness Consensus Committee. *Neuromodulation* 2014; **17**: 515–50; discussion 550.

12 Bikson M, Brunoni AR, Charvet LE, *et al.* Rigor and reproducibility in research with transcranial electrical stimulation: An NIMH-sponsored workshop. *Brain Stimul* 2018; **11**: 465–80.

13 Sandars M, Cloutman L, Woollams AM. Taking Sides: An Integrative Review of the Impact of Laterality and Polarity on Efficacy of Therapeutic Transcranial Direct Current Stimulation for Anomia in Chronic Poststroke Aphasia. *Neural Plast* 2016; **2016**: 8428256.

14 Parikh SV, Quilty LC, Ravitz P, *et al.* Canadian Network for Mood and Anxiety Treatments (CANMAT) 2016 Clinical Guidelines for the Management of Adults with Major Depressive Disorder: Section 2. Psychological Treatments. *Can J Psychiatry* 2016; **61**: 524–39.

15 Legatt AD, Emerson RG, Epstein CM, *et al.* ACNS Guideline: Transcranial Electrical Stimulation Motor Evoked Potential Monitoring. *J Clin Neurophysiol* 2016; **33**: 42–50.

16 Lefaucheur J-P, Antal A, Ayache SS, *et al.* Evidence-based guidelines on the therapeutic use of transcranial direct current stimulation (tDCS). *Clin Neurophysiol* 2017; **128**: 56–92.

17 Gillick BT, Gordon AM, Feyma T, *et al.* Non-Invasive Brain Stimulation in Children With Unilateral Cerebral Palsy: A Protocol and Risk Mitigation Guide. *Front Pediatr* 2018; **6**: 56.

18 Fregni F, El-Hagrassy MM, Pacheco-Barrios K, *et al.* Evidence-based guidelines and secondary meta-analysis for the use of transcranial direct current stimulation (tDCS) in neurological and psychiatric disorders. *Int J Neuropsychopharmacol* 2020; published online July 26. DOI:10.1093/ijnp/pyaa051.

19 Cruccu G, Garcia-Larrea L, Hansson P, *et al.* EAN guidelines on central neurostimulation therapy in chronic pain conditions. *Eur J Neurol* 2016; **23**: 1489–99.

20 Charvet LE, Shaw MT, Bikson M, Woods AJ, Knotkova H. Supervised transcranial direct current stimulation (tDCS) at home: A guide for clinical research and practice. *Brain Stimul* 2020; **13**: 686–93.

21 Charvet LE, Kasschau M, Datta A, *et al.* Remotely-supervised transcranial direct current stimulation (tDCS) for clinical trials: guidelines for technology and protocols. *Front Syst Neurosci* 2015; **9**: 26.

22 Bikson M, Hanlon CA, Woods AJ, *et al.* Guidelines for TMS/tES Clinical Services and Research through the COVID-19 Pandemic. DOI:10.31234/osf.io/82bmu.

23 Antal A, Alekseichuk I, Bikson M, *et al.* Low intensity transcranial electric stimulation: Safety, ethical, legal regulatory and application guidelines. *Clin Neurophysiol* 2017; **128**: 1774–809.

24 Shiozawa P, Cordeiro Q, Cho HJ, Trevizol AP, Brietzke E. A critical review of trials of transcranial direct current stimulation and trigeminal nerve stimulation for depression: the issue of treatment-emergent mania. *Trends Psychiatry Psychother* 2017; **39**: 48–53.

25 Santos FH, Mosbacher JA, Menghini D, Rubia K, Grabner RH, Cohen Kadosh R. Effects of transcranial stimulation in developmental neurocognitive disorders: A critical appraisal. *Prog Brain Res* 2021; **264**: 1–40.

26 Sanches C, Stengel C, Godard J, *et al.* Past, present, and future of non-invasive brain stimulation approaches to treat cognitive impairment in neurodegenerative diseases: Time for a comprehensive critical review. *Front Aging Neurosci* 2021; **12**: 578339.

27 McClintock SM, Kallioniemi E, Martin DM, Kim JU, Weisenbach SL, Abbott CC. A Critical Review and Synthesis of Clinical and Neurocognitive Effects of Noninvasive Neuromodulation Antidepressant Therapies. *Focus* 2019; **17**: 18–29.

28 Maatoug R, Bihan K, Duriez P, *et al.* Non-invasive and invasive brain stimulation in alcohol use disorders: A critical review of selected human evidence and methodological considerations to guide future research. *Compr Psychiatry* 2021; **109**: 152257.

29 Lucchiari C, Sala PM, Vanutelli ME. Promoting Creativity Through Transcranial Direct Current Stimulation (tDCS). A Critical Review. *Front Behav Neurosci* 2018; **12**: 167.

30 Godeiro C, França C, Carra RB, *et al.* Use of non-invasive stimulation in movement disorders: a critical review. *Arq Neuropsiquiatr* 2021; **79**: 630–46.

31 Cappon D, Jahanshahi M, Bisiacchi P. Value and Efficacy of Transcranial Direct Current Stimulation in the Cognitive Rehabilitation: A Critical Review Since 2000. *Front Neurosci* 2016; **10**: 157.

32 Workman CD, Fietsam AC, Rudroff T. Tolerability and Blinding of Transcranial Direct Current Stimulation in People with Parkinson's Disease: A Critical Review. *Brain Sci* 2020; **10**. DOI:10.3390/brainsci10070467.

33 ALHarbi MF, Armijo-Olivo S, Kim ES. Transcranial direct current stimulation (tDCS) to improve naming ability in post-stroke aphasia: A critical review. Behavioural Brain Research. 2017; **332**: 7–15.

34 Brunoni AR, Nitsche MA, Bolognini N, *et al.* Clinical research with transcranial direct current stimulation (tDCS): challenges and future directions. *Brain Stimul* 2012; **5**: 175–95.

35 Brunoni AR, Fregni F. Clinical trial design in non-invasive brain stimulation psychiatric research. *Int J Methods Psychiatr Res* 2011; **20**: e19–30.

36 Bikson M, Grossman P, Thomas C, *et al.* Safety of Transcranial Direct Current Stimulation: Evidence Based Update 2016. *Brain Stimul* 2016; **9**: 641–61.

37 Ekhtiari H, Tavakoli H, Addolorato G, *et al.* Transcranial electrical and magnetic stimulation (tES and TMS) for addiction medicine: A consensus paper on the present state of the science and the road ahead. *Neurosci Biobehav Rev* 2019; **104**: 118–40.

38 Martelletti P, Jensen RH, Antal A, *et al.* Neuromodulation of chronic headaches: position statement from the European Headache Federation. *J Headache Pain* 2013; **14**: 86.

39 Grimaldi G, Argyropoulos GP, Boehringer A, *et al.* Non-invasive Cerebellar Stimulation—a Consensus Paper. *Cerebellum* 2014; **13**: 121–38.

40 Buch ER, Santarnecchi E, Antal A, *et al.* Effects of tDCS on motor learning and memory formation: A consensus and critical position paper. *Clin Neurophysiol* 2017; **128**: 589–603.

41 Baptista AF, Fernandes AMBL, Sá KN, *et al.* Latin American and Caribbean consensus on noninvasive central nervous system neuromodulation for chronic pain management (LAC2-NIN-CP). *Pain Rep* 2019; **4**: e692.

42 Aparício LVM, Guarienti F, Razza LB, Carvalho AF, Fregni F, Brunoni AR. A Systematic Review on the Acceptability and Tolerability of Transcranial Direct Current Stimulation Treatment in Neuropsychiatry Trials. *Brain Stimul* 2016; **9**: 671–81.

43 Moffa AH, Brunoni AR, Fregni F, *et al.* Safety and acceptability of transcranial direct current stimulation for the acute treatment of major depressive episodes: Analysis of individual patient data. *J Affect Disord* 2017; **221**: 1–5.

44 Nikolin S, Huggins C, Martin D, Alonzo A, Loo CK. Safety of repeated sessions of transcranial direct current stimulation: A systematic review. *Brain Stimul* 2018; **11**: 278–88.

45 Brunoni AR, Amadera J, Berbel B, Volz MS, Rizzerio BG, Fregni F. A systematic review on reporting and assessment of adverse effects associated with transcranial direct current stimulation. *Int J Neuropsychopharmacol* 2011; **14**: 1133–45.

46 Carvalho AF, Sharma MS, Brunoni AR, Vieta E, Fava GA. The Safety, Tolerability and Risks Associated with the Use of Newer Generation Antidepressant Drugs: A Critical Review of the Literature. *Psychother Psychosom* 2016; **85**: 270–88.

47 Rosenblat JD, Kurdyak P, Cosci F, *et al.* Depression in the medically ill. *Aust N Z J Psychiatry* 2020; **54**: 346–66.

48 Woods AJ, Antal A, Bikson M, *et al.* A technical guide to tDCS, and related non-invasive brain stimulation tools. *Clin Neurophysiol* 2015; published online Nov 22. DOI:10.1016/j.clinph.2015.11.012.

49 Chu C-S, Li C-T, Brunoni AR, *et al.* Cognitive effects and acceptability of non-invasive brain stimulation on Alzheimer's disease and mild cognitive impairment: a component network meta-analysis. *J Neurol Neurosurg Psychiatry* 2021; **92**: 195–203.

50 Busch AB, Sugarman DE, Horvitz LE, Greenfield SF. Telemedicine for treating mental health and substance use disorders: reflections since the pandemic. *Neuropsychopharmacology* 2021; **46**: 1068–70.

51 Kopelovich SL, Monroe-DeVita M, Buck BE, *et al.* Community Mental Health Care Delivery During the COVID-19 Pandemic: Practical Strategies for Improving Care for People with Serious Mental Illness. *Community Ment Health J* 2021; **57**: 405–15.

52 Sathappan AV, Luber BM, Lisanby SH. The Dynamic Duo: Combining noninvasive brain stimulation with cognitive interventions. *Prog Neuropsychopharmacol Biol Psychiatry* 2019; **89**: 347–60.

53 Harris PA, Taylor R, Thielke R, Payne J, Gonzalez N, Conde JG. Research electronic data capture (REDCap)--a metadata-driven methodology and workflow process for providing translational research informatics support. *J Biomed Inform* 2009; **42**: 377–81.

54 Vaidyam AN, Linggonegoro D, Torous J. Changes to the Psychiatric Chatbot Landscape: A Systematic Review of Conversational Agents in Serious Mental Illness: Changements du paysage psychiatrique des chatbots: une revue systématique des agents conversationnels dans la maladie mentale sérieuse. *Can J Psychiatry* 2021; **66**: 339–48.

55 Li D. 5G and intelligence medicine-how the next generation of wireless technology will reconstruct healthcare? *Precis Clin Med* 2019; **2**: 205–8.

56 Torous J, Onnela J-P, Keshavan M. New dimensions and new tools to realize the potential of RDoC: digital phenotyping via smartphones and connected devices. *Transl Psychiatry* 2017; **7**: e1053.

57 Zanos S. Closed-Loop Neuromodulation in Physiological and Translational Research. *Cold Spring Harb Perspect Med* 2019; **9**. DOI:10.1101/cshperspect.a034314.

58 Polhemus AM, Novák J, Ferrao J, *et al.* Human-Centered Design Strategies for Device Selection in mHealth Programs: Development of a Novel Framework and Case Study. *JMIR Mhealth Uhealth* 2020; **8**: e16043.

59 Suen PJC, Doll S, Batistuzzo MC, *et al.* Association between tDCS computational modeling and clinical outcomes in depression: data from the ELECT-TDCS trial. *Eur Arch Psychiatry Clin Neurosci* 2021; **271**: 101–10.

60 Woods AJ, Bryant V, Sacchetti D, Gervits F, Hamilton R. Effects of Electrode Drift in Transcranial Direct Current Stimulation. *Brain Stimul* 2015; **8**: 515–9.

61 Hunold A, Ortega D, Schellhorn K, Haueisen J. Novel flexible cap for application of transcranial electrical stimulation: a usability study. *Biomed Eng Online* 2020; **19**: 50.

62 Pilloni G, Woods AJ, Charvet L. No risk of skin lesion or burn with transcranial direct current stimulation (tDCS) using standardized protocols. Brain Stimul. 2021; **14**: 511–2.

63 Goerigk SA, Padberg F, Bühner M, *et al.* Distinct trajectories of response to prefrontal tDCS in major depression: results from a 3-arm randomized controlled trial. *Neuropsychopharmacology* 2021; **46**: 774–82.

64 Steenbergen L, Sellaro R, Hommel B, Lindenberger U, Kuhn S, Colzato LS. 'Unfocus' on foc.us: commercial tDCS headset impairs working memory. *Exploratory Brain Research* 2015; published online Aug 18. DOI:10.1007/s00221-015-4391-9.

65 Antich-Isern P, Caro-Barri J, Aparicio-Blanco J. The combination of medical devices and medicinal products revisited from the new European legal framework. *Int J Pharm* 2021; : 120992.

66 Darrow JJ, Avorn J, Kesselheim AS. FDA Regulation and Approval of Medical Devices: 1976-2020. *JAMA* 2021; **326**: 420–32.

67 Wexler A. A pragmatic analysis of the regulation of consumer transcranial direct current stimulation (TDCS) devices in the United States. *J Law Biosci* 2015; **2**: 669–96.

68 Bikson M, Paneri B, Mourdoukoutas A, *et al.* Limited output transcranial electrical stimulation (LOTES-2017): Engineering principles, regulatory statutes, and industry standards for wellness, over-the-counter, or prescription devices with low risk. *Brain Stimul* 2018; **11**: 134–57.

69 Onodera R, Sengoku S. Innovation process of mHealth: An overview of FDA-approved mobile medical applications. *Int J Med Inform* 2018; **118**: 65–71.

70 Aljedaani B, Babar MA. Challenges With Developing Secure Mobile Health Applications: Systematic Review. *JMIR Mhealth Uhealth* 2021; **9**: e15654.

71 Valiengo L da CL, Goerigk S, Gordon PC, *et al.* Efficacy and Safety of Transcranial Direct Current Stimulation for Treating Negative Symptoms in Schizophrenia: A Randomized Clinical Trial. *JAMA Psychiatry* 2019; published online Oct 16. DOI:10.1001/jamapsychiatry.2019.3199.

72 Brunoni AR, Valiengo L, Baccaro A, *et al.* The sertraline vs. electrical current therapy for treating depression clinical study: results from a factorial, randomized, controlled trial. *JAMA Psychiatry* 2013; **70**: 383–91.

73 Brunoni AR, Moffa AH, Sampaio-Junior B, *et al.* Trial of Electrical Direct-Current Therapy versus Escitalopram for Depression. *N Engl J Med* 2017; **376**: 2523–33.

74 D'Urso G, Brunoni AR, Mazzaferro MP, Anastasia A, de Bartolomeis A, Mantovani A. Transcranial direct current stimulation for obsessive-compulsive disorder: A randomized, controlled, partial crossover trial. *Depress Anxiety* 2016; **33**: 1132–40.

75 Patsopoulos NA. A pragmatic view on pragmatic trials. *Dialogues Clin Neurosci* 2011; **13**: 217–24.

76 Inan OT, Tenaerts P, Prindiville SA, *et al.* Digitizing clinical trials. *NPJ Digit Med* 2020; **3**: 101.

77 Pallmann P, Bedding AW, Choodari-Oskooei B, *et al.* Adaptive designs in clinical trials: why use them, and how to run and report them. *BMC Med* 2018; **16**: 29.

78 Brunoni AR, Sampaio-Junior B, Moffa AH, *et al.* The Escitalopram versus Electric Current Therapy for Treating Depression Clinical Study (ELECT-TDCS): rationale and study design of a non-inferiority, triple-arm, placebo-controlled clinical trial. *Sao Paulo Med J* 2015; **133**: 252–63.

79 Valiengo L, Gordon PC, de Carvalho JB, *et al.* Schizophrenia TreAtment with electRic Transcranial Stimulation (STARTS): design, rationale and objectives of a randomized, double-blinded, sham-controlled trial. *Trends Psychiatry Psychother* 2019; **41**: 104–11.

80 Gunduz ME, Pacheco-Barrios K, Bonin Pinto C, *et al.* Effects of Combined and Alone Transcranial Motor Cortex Stimulation and Mirror Therapy in Phantom Limb Pain: A Randomized Factorial Trial. *Neurorehabil Neural Repair* 2021; **35**: 704–16.

81 Magill N, Knight R, McCrone P, Ismail K, Landau S. A scoping review of the problems and solutions associated with contamination in trials of complex interventions in mental health. *BMC Med Res Methodol* 2019; **19**: 4.

82 Razza LB, Afonso Dos Santos L, Borrione L, *et al.* Appraising the effectiveness of electrical and magnetic brain stimulation techniques in acute major depressive episodes: an umbrella review of meta-analyses of randomized controlled trials. *Braz J Psychiatry* 2020; published online Oct 23. DOI:10.1590/1516-4446-2020-1169.

83 Khadka N, Borges H, Paneri B, *et al.* Adaptive current tDCS up to 4 mA. *Brain Stimul* 2020; **13**: 69–79.

84 Fonteneau C, Mondino M, Arns M, *et al.* Sham tDCS: A hidden source of variability? Reflections for further blinded, controlled trials. *Brain Stimul* 2019; **12**: 668–73.

85 Turner C, Jackson C, Learmonth G. Is the ‘end-of-study guess’ a valid measure of sham blinding during transcranial direct current stimulation? *Eur J Neurosci* 2021; **53**: 1592–604.

86 Brunoni AR, Schestatsky P, Lotufo PA, Benseñor IM, Fregni F. Comparison of blinding effectiveness between sham tDCS and placebo sertraline in a 6-week major depression randomized clinical trial. *Clin Neurophysiol* 2014; **125**: 298–305.

87 Fassi L, Kadosh RC. Is it all in our head? When subjective beliefs about receiving an intervention are better predictors of experimental results than the intervention itself. bioRxiv. 2020; published online Dec 7. DOI:10.1101/2020.12.06.411850.

88 Insel TR. Digital Phenotyping: Technology for a New Science of Behavior. *JAMA* 2017; **318**: 1215–6.
